# Comparing implementation strategies for optimizing depression care: A randomized control trial

**DOI:** 10.1101/2025.01.14.25320533

**Authors:** Nathalie Moise, Maria Serafini, Danielle Rome, Jennifer Mizhquiri Barbecho, Kirali Genao, Siqin Ye, Andrea T. Duran, Joseph E. Schwartz

## Abstract

**Importance:** Less than a third of depressed primary care patients experience clinical improvement, in part due to a lack of focus on treatment optimization (e.g., intensification).

**Objective:** To compare the impact of implementation and behavioral science informed system and multi-level strategies on population-wide treatment optimization in integrated/collaborative care model (CoCM) settings.

**Design:** Comparative effectiveness randomized controlled trial

**Setting:** 5 Primary care clinics with a mature integrated/CoCM

**Participants:** 44 primary care physicians and their patients with elevated depressive symptoms eligible for treatment optimization

**Exposures:** System-level strategy (i.e., enhanced usual care [EUC]) focused on staff and behavioral health provider (BHP) activation vs. multi-level strategy (intervention) involving BHP activation, primary care provider (PCP) behavioral support and a patient activation/psychoeducation tool (DepCare)

**Main outcomes and measures:** Patient optimization (e.g., filling a new, intensified/augmented, or previously nonadherent antidepressant and/or completing a new integrated/CoCM visit) during the 4 months following an index visit and PCP optimization (e.g., placing a referral for any integrated/CoCM service and/or initiating, intensifying, switching and/or combining antidepressant medications) at an index visit. We used multilevel logistic regression analysis (level 1 is the patient with an eligible visit, level 2 the PCP) to test our hypotheses. Odds ratios (ORs) and 95% CIs were based on these analyses.

**Results:** There were 605 eligible patients with 757 visits in the post-implementation period. The mean age was 48 (SD=17); 486 (80%) were female, 15% Black, 51% Hispanic and 32% Spanish speaking; 41% were on an antidepressant. Patient treatment optimization in the intervention vs. EUC arms was 39.1% vs. 44.9% (OR=0.78; 95% CI 0.50, 1.22, p =0.27). Pre- vs. post-implementation, patient treatment optimization increased from 30.0% to 39.1% (p=0.10) and 30.4% to 44.9% (p=0.001) in the intervention and EUC arms (p=0.22 for differential change). There were similar trends in PCP optimization behaviors. There was low fidelity to the DepCare tool.

**Conclusions and relevance:** Our study demonstrates little added benefit of a multi-level over a system-level strategy as it relates to treatment optimization, with only system-level strategies demonstrating pre-post improvements. Negative unintended impacts of multi-level, particularly clinician targeted, strategies should be explored.

**Key Points:** *Question:* Is a theory-informed system-level strategy better than a multi-level strategy for improving population wide depression treatment optimization in integrated primary care settings?

*Findings:* In this comparative effectiveness randomized control trial of 2 implementation strategies for improving depression treatment optimization in integrated care settings, a multi-level strategy was no better than a system-level strategy for improving patient and clinician treatment optimization behaviors. Only the system-level strategy exhibited significant pre-post improvement in patient optimization.

*Meaning:* This is the first study to combine implementation and behavioral science to target treatment optimization in integrated care settings. We suggest that multi-level strategies that include clinician behavioral support may not be helpful and even harmful for improving population wide outcomes.

## Introduction

Only 27% of depressed primary care patients see significantly clinical improvement.^1^ This rate has remained unchanged for two decades,^1^ despite higher proportions of visits addressing mental health,^2^ primary care provider (PCP) comfort/knowledge,^3^ and implementation^4,5^ of integrated care programs, particularly collaborative care models (CoCM) involving collaboration between PCPs, behavioral health providers (BHP), and consulting psychiatrists proven to equitably improve depression outcomes.^6–9^ One reason effective treatments have less population-wide real world impact is suboptimal treatment optimization,^10,11^ which involves treatment intensification and improved reach of treat-to-target integrated/CoCM. However, few studies target treatment optimization, particularly in integrated care settings with systems in place to support access.^12^

A recent Lancet Commission noted the integral role of implementation science^13^ in improving mental health outcomes. These experts and others call for additional research on scaling and sustaining integrated care programs, testing theory-informed, multi-level strategies (e.g., patient, PCP, and/or system), and leveraging behavioral sciences.^14,15^ However, it remains unclear whether more intensive multi-level strategies, heralded as key to achieving health equity,^16^ are more effective than system-level strategies alone in promoting practice-wide treatment optimization in integrated care settings. Our study was informed by both behavioral and implementation science, specifically the Behavior Change Wheel^17^ – a transtheoretical framework for developing theory-informed, multi-level strategies. We aimed to compare the impact of a multi-level (aimed at systems/BHPs, PCPs, and patients) strategy (DepCare) (intervention) to a system-level strategy (EUC) aimed at care-manager practices on both patient (primary) and PCP (secondary) treatment optimization behaviors in socioeconomically diverse integrated care settings.

## Methods

### Study design

This was a PCP cluster randomized control trial (**Figure 1**). Analyses included all eligible patients of enrolled PCPs. To examine implementation outcomes and mechanisms by which strategies worked or didn’t work, a subset of patients and PCPs were consented and surveyed. Details of the strategy design and usability testing for TRANSFORM DepCare were previously described.^18,19^ The study protocol (Supplementary Material 1) was approved by the Institutional Review Board at Columbia University Irving Medical Center (CUIMC). The study adhered to Standards for Reporting Implementation Studies (StaRi Checklist). Informed by the Reach, Effectiveness, Adoption, Implementation and Maintenance (RE-AIM) framework (Supplementary Material 1),^20^ our study focused on patient (reach) and provider (adoption) optimization as well as strategy fidelity (implementation). The pre-implementation period took place from 7/31/2020 to 7/31/2021, implementation period 8/1/2021 to 12/31/2021 (e.g. start-up activities), and post-implementation period 1/1/2022 to 12/31/2022. The maximum follow-up period and implementation strategies continued until 6/30/2023.

**Figure 1.**
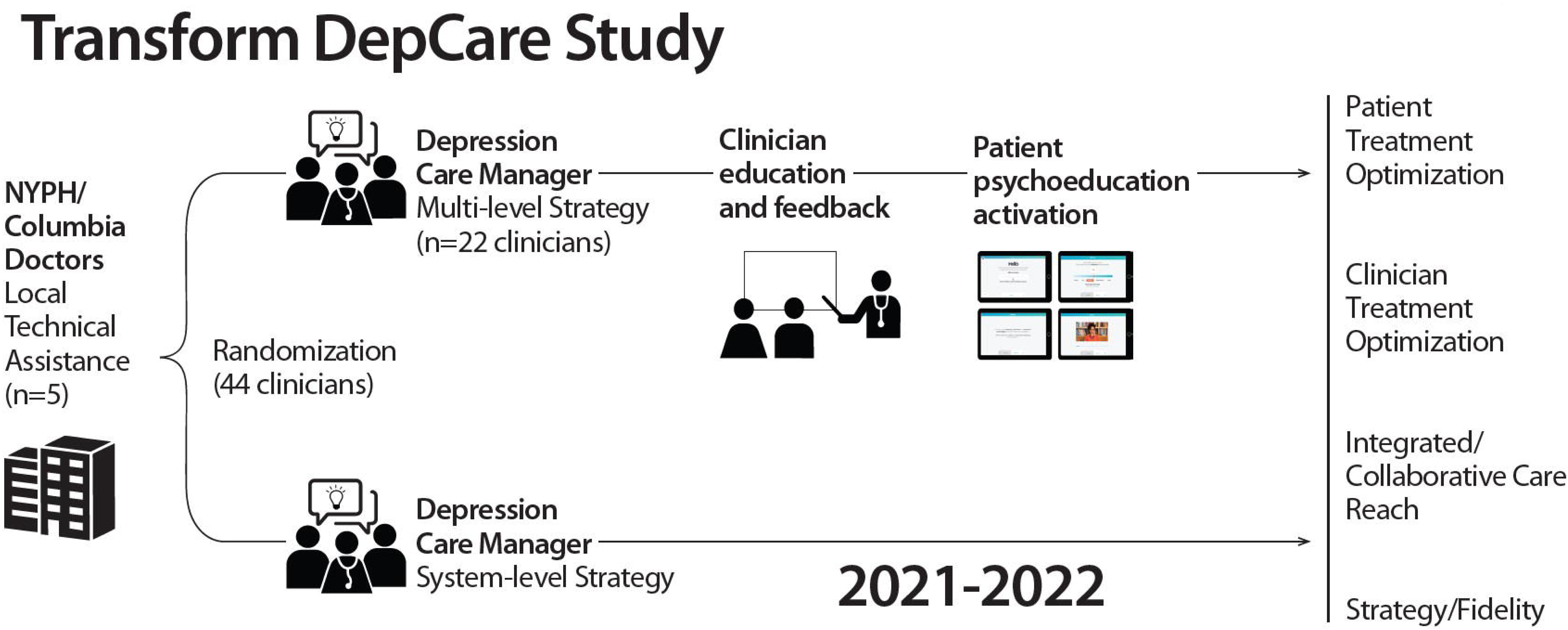
Design of the Transform DepCare Study. 44 primary care providers were randomized to receive either the system-level or multi-level strategy

### Study setting

Five CUIMC-affiliated primary care clinics with integrated/CoCM programs participated: 3 New York Presbyterian Ambulatory Care Network (ACN) clinics serving low-income, publicly insured, predominantly Hispanic and Black or African American patients and 2 Columbia Doctors Primary Care clinics serving diverse, commercially insured patients. Internal medicine physicians, nurse practitioners and/or graduate medical education trainees staffed clinics. All clinics had access to core components of reimbursable integrated/CoCM programs,^21^ including systematic depression and anxiety screening and integrated BHPs providing short-term measurement-based treatment with varied age and external support (**Supplementary Material 1**). All BHPs had low caseloads and were available for referrals.

### Eligibility

We included all PCPs serving adult patients in the 5 integrated/CoCM clinics whose medical directors agreed to participate in the trial (**Figure 2**). We identified patients via the electronic health record (EHR; Epic) with ≥1 completed visit with an enrolled PCP during the relevant study periods, hereafter referred to as index visits. We included patients ≥18 years old, English or Spanish speaking, and with elevated depressive symptoms (Patient Health Questionnaire [PHQ]-9≥10 and/or PHQ9≥5 and Generalized Anxiety Disorder [GAD]≥10) 0-14 days prior to index visits (e.g., during pre-visit e-check-ins). We excluded patients (based on International Classification of Disease-10 codes) with a diagnosis of coronary heart disease, dementia, intellectual development disorder, severe mental illness, substance abuse or pregnancy as well as a documented PHQ9 <10 or PHQ ≥5 and GAD <10. We then excluded patients actively enrolled (i.e., seen in prior 3 months) in integrated care, the majority of which were CoCM but also included embedded psychiatry/behavioral/mental health programs already providing treatment optimization (**Supplementary Material 1**). Each visit with an eligible patient represented an opportunity to optimize treatment and patients could be eligible at ≥1 visit (**eTable 1, Supplementary Material 2**). Our pragmatic approach meant patients could have outside psychiatrists and/or therapists not captured in Epic but aligned with our aim to improve optimization regardless of baseline treatment. Once patients filled an antidepressant or completed an integrated/CoCM visit, subsequent “index visits” were excluded (**Figure 2**). Eligibility was confirmed by self-report for the subgroup of consented patients.

**Figure 2.** Consort Diagram. Flow of the of eligibility criteria for the post-implementation patients.

### Recruitment, Randomization and Allocation

We stratified PCPs at the 5 participating practices by training level (resident, chief/attending, NP). In 7/2021, we used a computer-generated allocation table incorporating permuted block randomization within each stratum (created by the study’s blinded biostatistician [JES]) to assign 36 PCPs to either the intervention (i.e., multi-level strategy) or enhanced usual care [EUC] (i.e., system-level strategy) arm. Eight new PCPs (e.g., incoming residents, Columbia Doctors PCPs) were randomly allocated prior to trial initiation for a total of 44 PCPs.

### Intervention and EUC Arms

We previously described the theoretical underpinnings, strategy design, and usability testing of our multi-component, multi-level strategy (**Supplementary Material 1**).^18,19^ Informed by stakeholder interviews with mature (>2 years) integrated/CoCM^22^ as well as the BCW^17^ and Dynamic Sustainability Framework,^23^ we mapped barriers to behavioral change techniques. While initially focusing on patient-level strategies, system and multi-level strategies emerged. A multidisciplinary team then iteratively designed, user-tested and adapted disseminable strategies to target optimization behaviors and address emerging COVID-19 driven contextual factors.^18,19^

#### EUC arm

We delivered the system-level strategy as part of a quality improvement initiative that included (1) a depression screening marketing and education video disseminated to staff (2) navigator-assisted pre-visit depression and anxiety screening and (3) quarterly local technical assistance (TA) to activate/problem solve integrated/CoCM programs with BHPs, administrators, and PCP champions. PCPs (and their patients) in this EUC arm received no direct intervention.

#### Intervention arm

In addition to the system-level strategy above, PCPs in the intervention arm received (1) a motivational and marketing video promoting CoCM and treatment optimization (2) access to a treatment optimization smartphrase in Epic and (3) regular emailed motivational and audit/feedback newsletters (4) EHR delivered decision support among the subset of eligible patients consenting to receive our DepCare tool (which provided symptom recognition support, psychoeducation, activation videos promoting treatment engagement and personalized medication selection support). Following COVID-19 disruptions, the IRB required patients’ clinical teams to gauge interest and obtain preliminary consent for tool delivery (in lieu of planned dissemination to all eligible patients) (**Supplementary Material 1**).

### Outcomes

The primary outcome was patient treatment optimization defined as initiating, intensifying, newly adhering to (if not filling 4 months prior) or augmenting antidepressants and/or completing an integrated/CoCM visit) during the 4 months following an index visit, regardless of whether the patient was in treatment at baseline. For patients with multiple index visits, we attributed the outcome to the most recent index visit. Prespecified secondary outcomes included (1) PCP treatment optimization at the index visit (i.e., [a] placing an integrated/CoCM care referral, [b] initiating, intensifying, switching and/or combining antidepressant medications and/or [c] documenting depression management/adherence counseling for those receiving DepCare tool); (2a) ≥2 integrated/CoCM visits and (2b) ≥2 antidepressant fills both at 6 months among those who met 4 month optimization definitions.

### Data Collection

Data were available via a CUIMC Data Warehouse query for referral orders, visits, flowsheets, and medication fill dates. Columbia Doctors did not use order-sets and referrals (only not patient behavior) were manually extracted via PCP notes/messages to BHPs. Among the subset of consented patients, two medically trained abstractors (DR, NM) blinded to group assignment independently reviewed the EHR for evidence of patient- and PCP-level optimization (using notes to identify impact on mental health visits outside CUIMC). Discrepancies were resolved through consensus. At the end of the study, we approached both intervention and EUC PCPs to complete an exit survey that included single-item burnout measures,^24^ net promoter score,^25^ and implementation outcomes related to the acceptability, feasibility, and appropriateness^26^ of integrated/CoCM. We measured fidelity by meeting attendance and newsletter/tool read/use/receipt.

### Power Calculation

Sample size calculation was informed by the observed effect sizes of our systematic review of multi-level, multi-component engagement (vs. EUC) trials in primary care settings as well as patient-preference/engagement interventions (vs. usual care), both of which showed effect sizes of about 20%.^27^ Allowing for clustering within PCPs (assuming an intraclass correlation [ICC]^28,29^ of 0.076 based on our preliminary data) and using the standard power calculation formula for group RCTs,^30^ we determined that 36 providers (18 per arm) with an average of 9 patients with eligible index visits/provider (N=324 patients in total) would provide 80% power to detect the hypothesized 20% difference between arms in patient optimization rates. In the actual study, we were able to randomize 44 providers (605 patients with ≥1 eligible index visit) and therefore had more than adequate power to detect an effect size of 20%. We were not powered to assess the patient-level strategy (**Supplementary Material 1**).

### Statistical analysis

We described baseline characteristics of categorical variables (frequencies and percentages) and quantitative variables (means [SDs]). We estimated a multilevel logistic regression model,^31,32^ where index visits (level 1) were nested within patients (level 2), nested within PCPs (level 3), to predict primary and secondary patient optimization behaviors (yes/no) with arm (intervention vs EUC) × Period (pre- vs. post-implementation) interaction (4 categories) as the only predictor.

We computed estimates of pre-specified contrasts for each outcome, tested using the Wald F-test (2-tailed, α=0.05) and obtained odds ratios (ORs) and their 95% CIs by exponentiating the contrast estimates and their 95% CIs. Patients’ outcomes were all analyzed in the arm to which their providers were randomized (intent-to-treat principle), regardless of whether they consented to the DepCare tool. For analyses of provider behaviors, we determined the number of patient index visits and proportion of patient index visits in which the provider optimized treatment, using these to estimate a multilevel binomial regression model predicting the proportion of index visits for which providers optimized. Missing data were treated as “non-optimized” behavior.

We performed exploratory, stratified analyses to determine effect differences in Columbia Doctors vs. ACN clinics. We further assessed the subgroup of patients consented to receive the Depcare tool vs. usual care. Sensitivity analyses excluded PCP champions involved in our system-level intervention. All analyses were performed using the GLIMMIX procedure in SAS 9.4.

## Results

Overall, there were 605 patients with 757 visits in the post-implementation period (325 patients with 389 visits in intervention arm vs. 295 patients with 368 visits in the EUC arm; 15 patients saw PCPs in both arms). The mean age was 48 (SD=17); 486 (80%) were female, 15% Black, 35% multi-racial; 51% Hispanic and 32% preferred English; 41% were on an antidepressant at their first index visit and 49% received care in the ACN (**Table 2**). Intervention patients were more likely to be older, historically marginalized, and non-English speaking in the ACN. In the pre-implementation period, there were 382 patients with 529 visits with few significant Period × Condition interactions in demographics (**eFigure 1, eTable 2, Supplementary Material 2**).

**Table 1.**
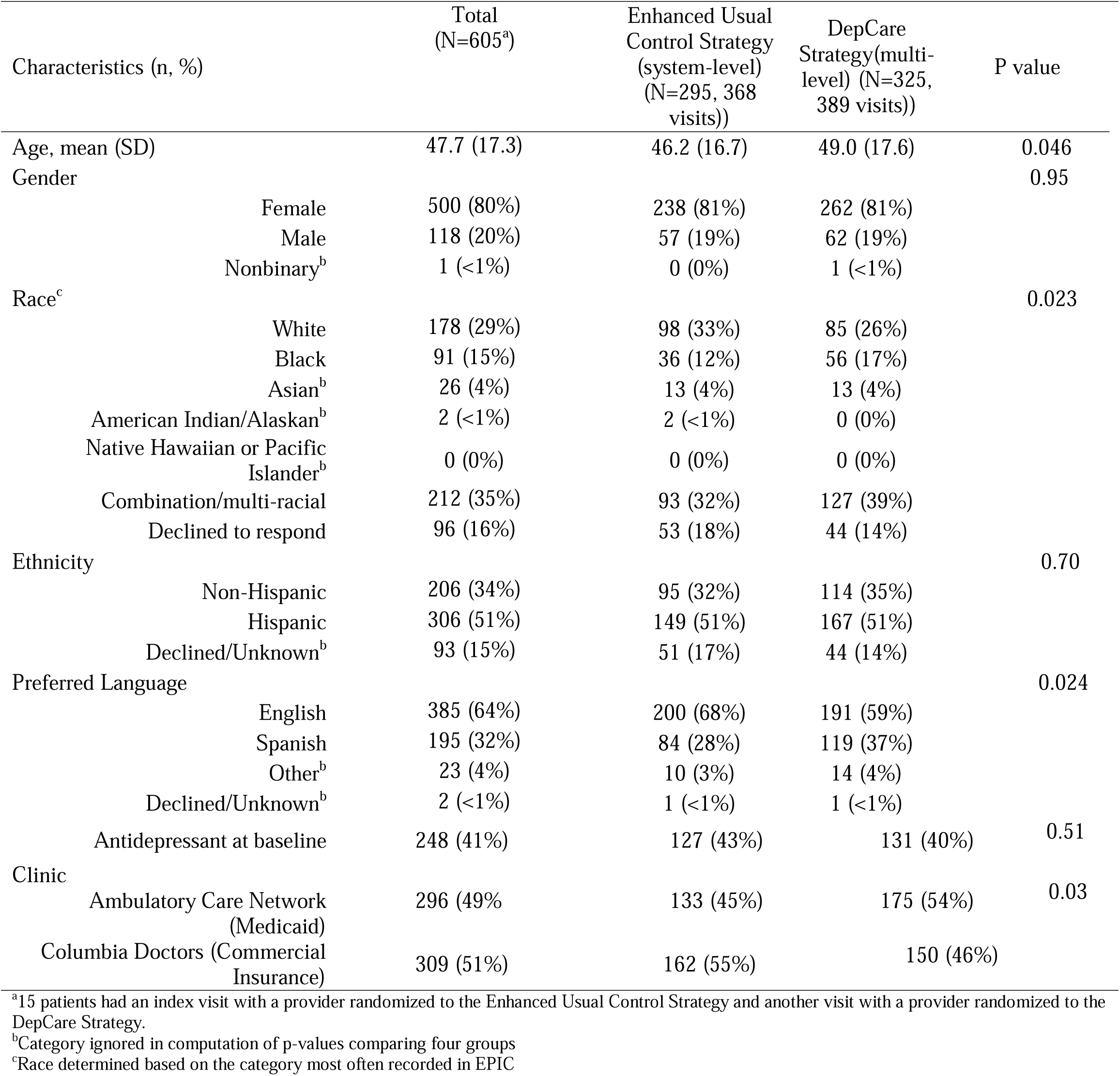
Patient Characteristics in the Post Strategy Period.

| Characteristics (n, %) |  | Total<br>(N=605<sup>a</sup>) | Enhanced Usual<br>Control Strategy<br>(system-level)<br>(N=295, 368<br>visits)) | DepCare<br>Strategy(multi-level) (N=325,<br>389 visits)) | P value |
| --- | --- | --- | --- | --- | --- |
| Age, mean (SD) |  | 47.7 (17.3) | 46.2 (16.7) | 49.0 (17.6) | 0.046 |
| Gender |  |  |  |  | 0.95 |
|  | Female | 500 (80%) | 238 (81%) | 262 (81%) |  |
|  | Male | 118 (20%) | 57 (19%) | 62 (19%) |  |
|  | Nonbinary<sup>b</sup> | 1 (<1%) | 0 (0%) | 1 (<1%) |  |
| Race<sup>c</sup> |  |  |  |  | 0.023 |
|  | White | 178 (29%) | 98 (33%) | 85 (26%) |  |
|  | Black | 91 (15%) | 36 (12%) | 56 (17%) |  |
|  | Asian<sup>b</sup> | 26 (4%) | 13 (4%) | 13 (4%) |  |
|  | American Indian/Alaskan<sup>b</sup> | 2 (<1%) | 2 (<1%) | 0 (0%) |  |
|  | Native Hawaiian or Pacific<br>Islander<sup>b</sup> | 0 (0%) | 0 (0%) | 0 (0%) |  |
|  | Combination/multi-racial | 212 (35%) | 93 (32%) | 127 (39%) |  |
|  | Declined to respond | 96 (16%) | 53 (18%) | 44 (14%) |  |
| Ethnicity |  |  |  |  | 0.70 |
|  | Non-Hispanic | 206 (34%) | 95 (32%) | 114 (35%) |  |
|  | Hispanic | 306 (51%) | 149 (51%) | 167 (51%) |  |
|  | Declined/Unknown<sup>b</sup> | 93 (15%) | 51 (17%) | 44 (14%) |  |
| Preferred Language |  |  |  |  | 0.024 |
|  | English | 385 (64%) | 200 (68%) | 191 (59%) |  |
|  | Spanish | 195 (32%) | 84 (28%) | 119 (37%) |  |
|  | Other<sup>b</sup> | 23 (4%) | 10 (3%) | 14 (4%) |  |
|  | Declined/Unknown<sup>b</sup> | 2 (<1%) | 1 (<1%) | 1 (<1%) |  |
| Antidepressant at baseline |  | 248 (41%) | 127 (43%) | 131 (40%) | 0.51 |
| Clinic |  |  |  |  | 0.03 |
|  | Ambulatory Care Network<br>(Medicaid) | 296 (49%) | 133 (45%) | 175 (54%) |  |
|  | Columbia Doctors (Commercial<br>Insurance) | 309 (51%) | 162 (55%) | 150 (46%) |  |

<sup>a</sup>15 patients had an index visit with a provider randomized to the Enhanced Usual Control Strategy and another visit with a provider randomized to the DepCare Strategy.
<sup>b</sup>Category ignored in computation of p-values comparing four groups
<sup>c</sup>Race determined based on the category most often recorded in EPIC

**Table 2.** Proportion of Eligible Visits with patient and provider optimization in the Post-Implementation Phase.

| **Outcomes**<br>N (%) of Eligible Visits<sup>a,b</sup> | **DepCare**<br>**Strategy Arm**<br>(n= 325 patients)<br>Yes:No (%) | **EUC Arm**<br>(n= 295 patients)<br>Yes:No (%) | **OR (95% CI)** | **P value** |
| --- | --- | --- | --- | --- |
| **Patient Optimization** |  |  |  |  |
| Any optimization | 132:206 (39.1%) | 136:167 (44.9%) | 0.78 (0.49, 1.22) | 0.27 |
| Integrated/CoCM Visit | 62:322 of (16.1%) | 67:299 (18.3%) | 0.95 (0.57, 1.58) | 0.85 |
| Any medication fill | 99:266 (27.1%) | 117:215 (35.2%) | 0.62 (0.40, 0.98) | 0.04 |
| **Primary Care Provider**<br>**Optimization** |  |  |  |  |
| Any optimization | 135:198 (40.5%) | 140:164 (42.1%) | 0.74 (0.42, 1.31) | 0.29 |
| Integrated/CoCM referral | 98:274 (26.3%) | 98:245 (28.6%) | 0.84 (0.51, 1.39) | 0.49 |
| Medication Intensification | 69:294 (19.0%) | 87:250 (25.8%) | 0.59 (0.30, 1.15) | 0.12 |
| **Any ≥2 antidepressant fills**<br>**and/or ≥2**<br>**integrated/CoCM visits** | 106:232 (31.4%) | 113:190 (37.3%) | 0.77 (0.50, 1.19) | 0.23 |
| **≥2 integrated/CoCM visits**<br>**over 6 months** | 39:345 (10.2%) | 56:310 (15.3%) | 0.73 (0.40, 1.30) | 0.28 |
| **≥2 antidepressant fills over**<br>**6 months** | 82:283 (22.5%) | 93:239 (28.0%) | 0.70 (0.46, 1.06) | 0.09 |

<sup>a</sup>Subsequent eligible index visit dropped if patient enrolled in integrated/collaborative care model or started a medication (i.e., each there were different number of eligible visits for each outcome).
<sup>b</sup> If an outcome occurred in period for >1 visit, optimization was allocated to the most recent visit

Treatment optimization among patients of intervention PCPs was 39.1% (16.1% integrated/CoCM visits, 27.1% antidepressants) vs. 44.9% (18.3% integrated/CoCM visits, 35.2% antidepressants) in the EUC arm (OR=0.78; 95% CI 0.49, 1.22, p =0.27). In the intervention vs. EUC arms, the proportion of eligible patients who had ≥2 integrated/CoCM visits and/or ≥2 prescription fills at 6 months was 31.4% (10.2% with ≥2 integrated/CoCM visits and 22.5% with ≥2 antidepressant fills) vs. 37.3% (15.3% with ≥2 integrated/CoCM visits and 28.0% with ≥2 antidepressant fills) (OR=0.77; 95% CI 0.50, 1.19, p =0.23). Intervention vs EUC PCP optimization was 40.5% (26.3%, integrated/CoCM referral, 19.0% antidepressant change/augmentation) vs. 46.1% (28.6% integrated/CoCM referral, 25.8% antidepressant change/augmentation) (OR=0.74; 95% CI 0.42, 1.31, p =0.29) (**Table 2**).

Pre-implementation vs. post-implementation, patient treatment optimization increased from 30.0% (9.5% ≥1 integrated/CC care visit, 23.9% ≥1 antidepressant fill) to 39.1% (16.1% for ≥1 integrated/CoCM visit, 27.1% ≥1 antidepressant fill) in the intervention arm (p=0.10) vs. 30.4% (10.5% ≥1 integrated/CoCM visit, 25.3% ≥1 antidepressant fill) to 44.9% (18.3% ≥1 integrated/CoCM visit, 35.2% for ≥1 antidepressant fill) in the EUC arm (p=0.001) (OR=0.71, 95% CI 0.40, 1.22; differential p=0.22). There were similar trends in PCP optimization behaviors (**Figure 3, eTable 3 Supplementary Material 2).**

**Figure 3.** Patient and provider optimization rates pre vs. post implementation.

Results were similar (1) excluding PCP champions and (2) in Columbia Doctors and ACN (older, historically marginalized) sites, though the former saw larger pre-post improvements **(eTables 4 and 5 Supplementary Material 2).**

### Implementation outcomes

Relevant to both arms (system-level strategy), ≥1 BHP/administrator from all 5 clinics attended all TA meetings (100%). Key TA discussions involved relapse prevention, PCP education, feedback on screening/optimization rates, and emerging CoCM best practices. In the intervention arm, 72.7% of PCPs read ≥1 activation email, 0% used the smartphrase, and 50.0% received ≥1 preference report; of 1865 patients receiving navigator-assisted screening, 62 patients were eligible, interested, referred by clinical team and consented to be randomized to receive the DepCare Tool (**eFigure 2, Supplementary Material 2**).

We approached all 44 PCPs, including 11 (25%) who retired/left/graduated, leaving 19 (43.2%; 7 intervention, 12 EUC) complete surveys; average age 36 years old; 61% were female; 50.0% endorsed burnout. Intervention vs. EUC PCPs found [range 1-5] integrated/CoCM acceptable (4.43 vs. 4.69), feasible (4.18 vs. 4.21) and appropriate (4.54 vs. 4.58) with a net promotor score of 9.06 (scale 0-10) (**eFigure 3, Supplementary Material 2**).

### Post Hoc Analyses

Of 62 consented patients, 5 were pilot participants and 1 withdrew consent (eFigure 3 Supplementary Material 2). Of 56 remaining patients (37 intervention, 19 EUC), a greater proportion of intervention vs. EUC arm patients optimized treatment (51.4% vs. 36.8%) including integrated/CoCM enrollment (27.0% vs. 10.5%) and antidepressant fill (32.4% vs. 15.8%); PCP optimization was 40.5% vs. 31.6%.

## Discussion

We leveraged implementation and behavioral science methods to develop and compare system and multi-level strategies in socio-economically diverse integrated/CoCM settings. We found no differences in patient and PCP treatment optimization, with significant pre-post improvements in the system-level only. Post hoc analysis found higher rates of treatment optimization among patients receiving the DepCare tool compared to EUC arm.

There have been calls to elucidate what strategies do or do not work in implementing and sustaining evidence-based mental health interventions.^14^ Integrated/CoCMs improve depressive symptoms, treatment initiation/access, and disparities.^9,33–37^ There is consensus that real world success similar to original trials requires intensive implementation support (e.g., TA, financing).^5,38–45^ Despite ongoing support, integrated/CoCM face declining caseloads over time due to waning resources and patient/PCP engagement.^22^ However, few studies focus on *sustainability*.

Our results suggest that once integrated/CoCMs are implemented, more intensive multi-level strategies targeting PCPs have little added benefit over system-level strategies, perhaps due to learned helplessness that develops from treatment relapse/resistance, reduced real-world effectiveness,^10^ and time constraints. Integrated/CoCM success hinges on PCP involvement/champions (e.g., to titrate antidepressants, conduct warm handoffs).^46^ The COVID-19 pandemic prompted increased attention to PCP burnout and moral injury.^47,48^ In fact, 30% of our PCPs left the practice. While PCPs in both arms found integrated/CoCM acceptable and feasible, it may be that visit asynchronous feedback/education highlighting PCP requirements caused more harm than good given lack of even system-level effects in the multi-level arm. Our post hoc analysis suggests that even real time feedback had limited impact on immediate PCP behavior, supporting evidence that PCPs ignore best practice alerts.^49^ EHR-embedded medication titration algorithms/protocols may be better suited to improving PCP optimization rates.^50^ Policymakers should consider statutes that limit centralized, automated referrals, such as the Centers for Medicare and Medicaid Services requirements that PCPs document patient consent prior to CoCM enrollment.^51^

Our significant pre-post findings in the EUC arm suggest that system-level strategies that activate BHPs may impact patient and marginally PCP behaviors, perhaps due to improved PCP communication, training, and patient outreach. Our findings highlight the importance of quality improvement efforts^44^ and limited added benefit of more complex strategies.^45^ We also provide insights into active ingredients (i.e., behavior change techniques like feedback on optimization behaviors)^19^ and disseminable strategy components. While we found no subgroup differences, pre-post improvements in optimization may have been driven by higher income sites, supporting data on low referral/follow-up rates in screen-detected historically marginalized patients.^11^ Finally, while widely recommended,^14,16^ multi-component and/or multi-level strategies are often compared to usual care, limiting conclusions about active or detrimental ingredients. Our comparative effectiveness design uncovered limitations of PCP-level strategies while signaling the potential of direct-to-patient marketing and activation tools,^52^ particularly those that address unique contextual and personal experiences, and impact both integrated and external help-seeking behaviors. Clinics are now deploying DepCare videos to waiting room TVs, and future research should explore other avenues for automatically delivering activation tools via Epic though our historically marginalized population necessitated technical assistance and lacked portals access.

Our trial had several limitations. There was moderate multi-level strategy fidelity (60-70%) and some components (i.e., newsletters) intensified over time, contributing to null or even delayed effects. Despite randomization, the intervention arm was older and historically marginalized though subgroup analyses were unrevealing. While supporting generalizability, CoCMs were heterogeneous, which along with time effects may have contributed to pre-post improvements particularly in high-income clinics, though we only saw improvements in the EUC arm and found no improvements in integrated/CoCM settings across NY State during this time (*manuscript*). Despite better real-world implications of relying on existing clinical staff to administer the DepCare tool, COVID-19/IRB stipulations precluded widespread dissemination, contributing to low reach. It is also possible that patients received multiple referrals at multiple eligible visits by PCPs though patients could only be optimized once; most had 1 visit and <3% were seen in both arms. Missing data was treated as “non-optimized” though absence or existence of visits/medications are reliable. cli

Overall, studies rarely compare system to multi-level strategies. We found little added benefit to our multi-level strategy, with significant pre-post improvements in the system-level arm only. Future work will examine CoCM fidelity, contextual factors, and underlying mechanisms using our survey data. Whether mental health behaviors are particularly resistant to PCP feedback compared to other diseases should be explored (e.g., PCP behavioral support improves hypertension outcomes).^53^ Additional research will need to examine the effect of direct-to-patient strategies.

## Data Availability

Data available: Yes
Data types: Deidentified participant data, Data dictionary
How to access data:
When available: With publication
Supporting Documents
Document types: Statistical/analytic code
How to access documents: >
When available: With publication
Additional Information
Who can access the data: Researchers whose proposed use of the data is approved.
Types of analyses: For a specified purpose
Mechanisms of data availability: With investigator support
Any additional restrictions: None

## Acknowledgments

The authors would like to thank Drs. Ian Kronish and Susan Essock for their critical revisions of this work. Thank you to Gennesis Zuleta for study recruitment and data processing. Thank you to Darlene Straussman for administrative support and the DepCare Advisory Board for study.

## Ethics approval and consent to participate

This study was approved by the Columbia University Institutional Review Board (study ID: AAAT6753).

## Consent for publication

Not applicable. Only aggregate patient-level data were used in this study.

## Availability of data and materials

The senior author (NM) and biostatistician (JES) had full access to the data and took responsibility for the integrity of the data and the accuracy of the data analysis. The datasets used and/or analyzed in this study will be made available in Open Science, https://www.cos.io upon request.

## Authors’ contributions

NM wrote the first draft of the manuscript. JES cleaned, managed, analyzed, and interpreted the data. DR and MS managed database and analyzed data. JBM managed the entire study. JES and NM drafted the data analysis and results sections. NM conceptualized the project, obtained funding and was a key contributor in writing the manuscript. KG and GZ collected and cleaned the data. All authors contributed to critical revision of the manuscript. All authors read and approved the final manuscript.

