## Supplementary material for "Comparing implementation strategies for optimizing depression care: A randomized control trial": Figure 2

**Figure 2. CONSORT Flow Diagram.** Enrollment, eligibility and analyses of post-implementation participants in the Transform DepCare Study

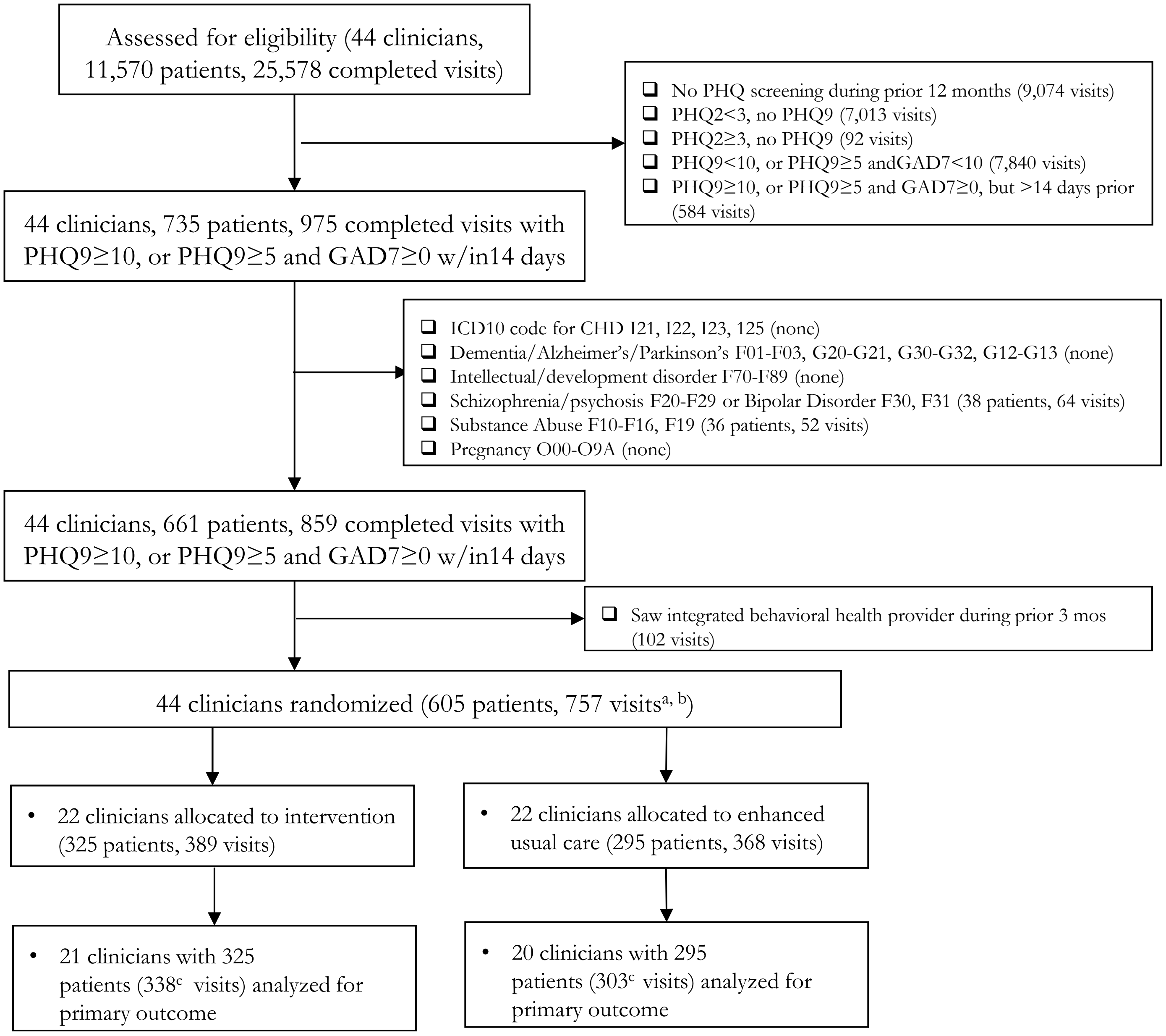


[a] Patients could have more than 1 visits, which ranged from 1-8/patient.

[b] 15 patients had visits with clinicians in both treatment conditions

[c] Represents number of index visits included in the analysis of the primary outcome. Number of visits varied for each outcome.
