## Supplementary material for "Comparing implementation strategies for optimizing depression care: A randomized control trial": Figure 3

**Figure 3. Pre-Post Patient and Provider Optimization Behaviors in the EUC vs. Intervention Arms of the Transform DepCare Study**

a

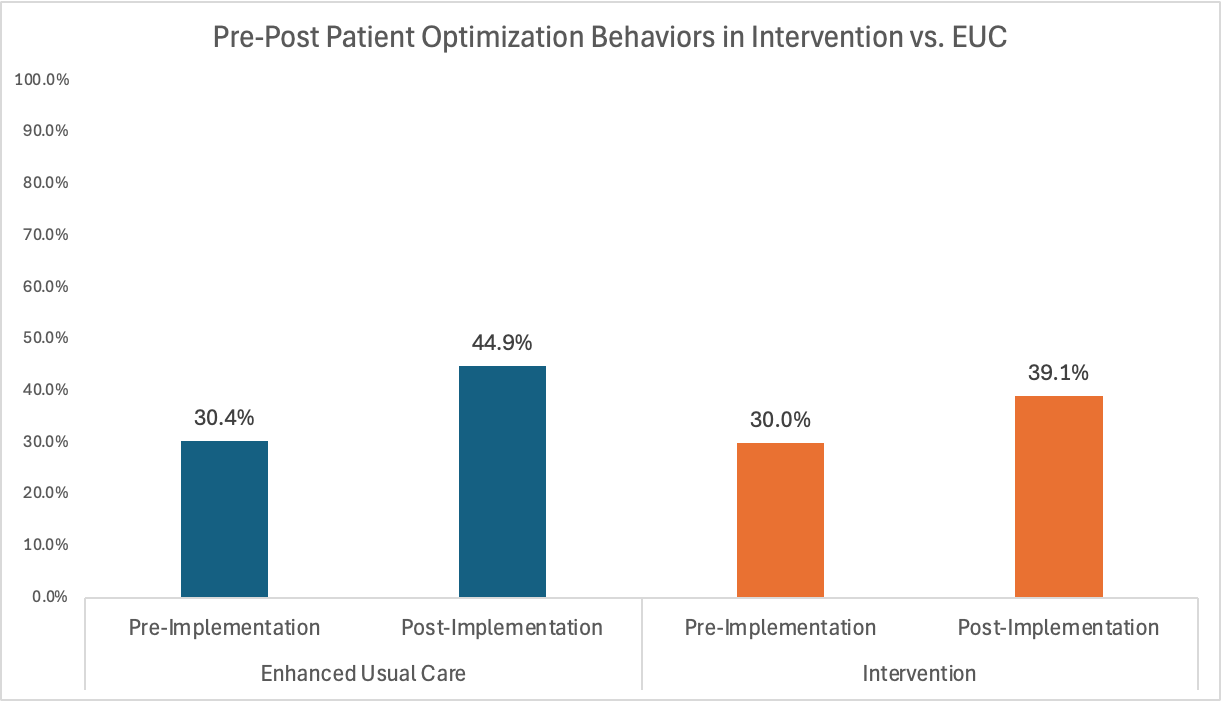

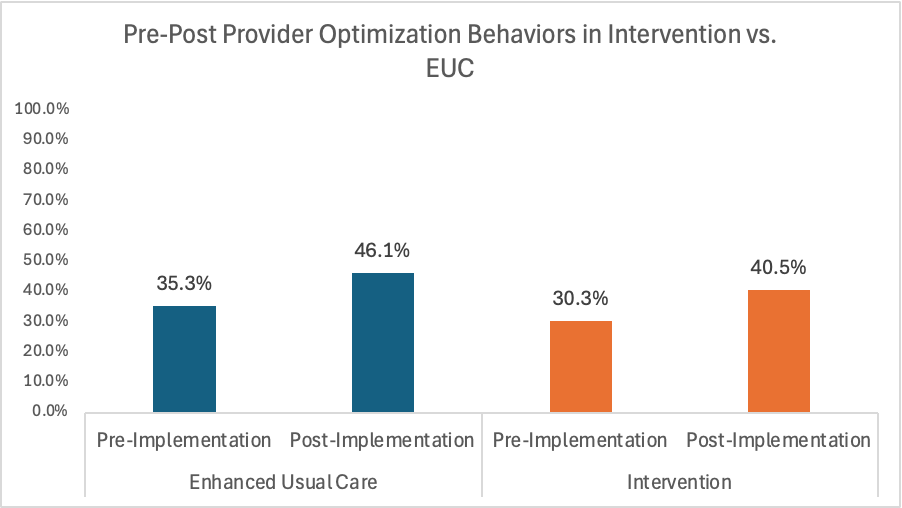

b

**

** Signifies p value of 0.001

^a^Differential p value = 0.22

^b^EUC p value = 0.05; Differential p value =0.52
