## Supplemental Material 1 for "Comparing implementation strategies for optimizing depression care: A randomized control trial"

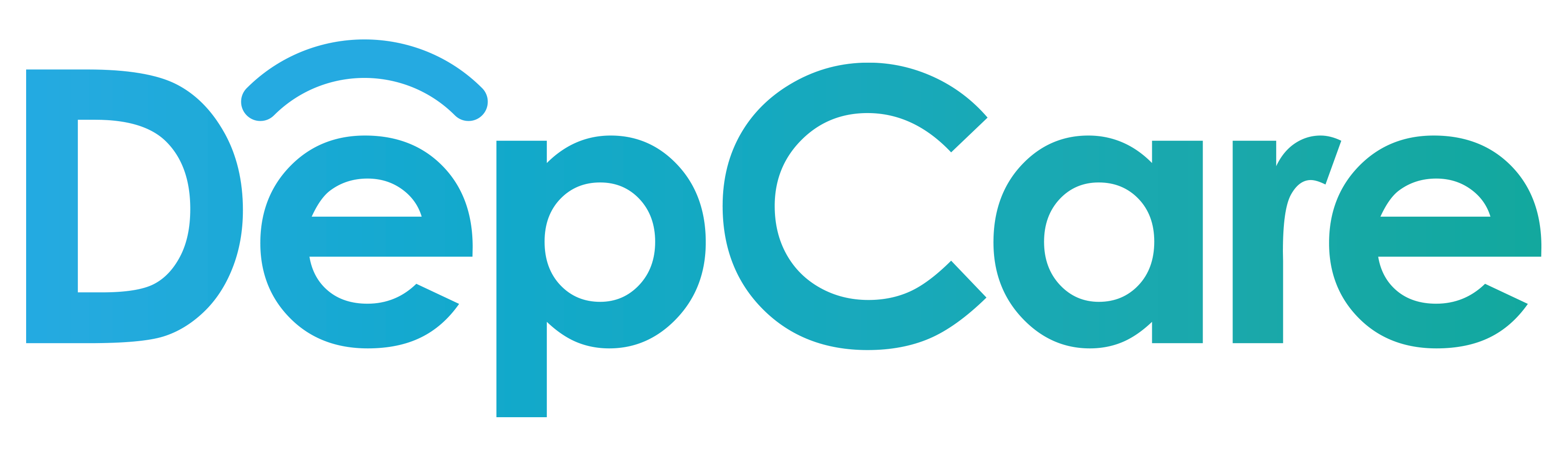

STUDY PROTOCOL
(Manual of Operations)

### Protocol Amendments

#### Summary of changes

| June 2020  [prior to trial launch] | Adapted intervention for post-COVID19 needs (e.g., tool refinement for remote vs. waiting room delivery as proposed), multi-level intervention in lieu of a patient-level intervention only |
| --- | --- |
| August 2021  [prior to trial launch] | Study design finalized for provider level cluster randomized control trial of system level intervention vs. multi-level intervention based on finalized implementation mapping, and COVID19 restrictions. Provider randomization completed. Stepped wedge design proposed in original aims of the grant deemed infeasible on discussion with statistician and study team members given clinics merged or co-located during COVID19. Following COVID19 disruptions, the IRB stipulated that members of the patient’s clinical team approach patients first to gauge interest, obtain preliminary consent, and refer to a research team member to receive the DepCare tool (in lieu of disseminating the tool to all eligible patients as planned). |
| October 2021  [Prior to trial launch]  October 2021  [prior to trial launch]  April 2022 | Primary outcomes relevant to patient and provider treatment optimization and sample size calculation completed. Clinicaltrials.gov submitted.  History of suicide attempt or ideations not included given not an exclusion criteria for CoCM and infeasible to exclude using EPIC (this remained via self-report for subgroup of consented patients)  First Technical Assistance Meeting conducted. To maintain blinding intervention providers were not invited to problem solving meetings (control providers were never slated to participate in problem solving meetings). Intervention PCPs received ongoing newsletters in lieu of meetings. |
| March 2024  March 2024  March 2024 | Adherence counseling component of provider optimization (secondary outcome) deemed infeasible to operationalize using electronic health record (EHR) data.  % of patients receiving any treatment at follow up deemed too similar to patient treatment optimization; baseline treatment rates reported instead.  At the time our grant was written and IRB submitted, we included some ICD9 codes, but focused on ICD10 codes only at the time of analyses given availability in our EHR. Given difficulty operationalizing using searchable field in the EHR, upcoming psychiatrist/care manager visit was removed from exclusion criteria as well as access to the internet/smart phone/planning to attend in person visit from inclusion criteria (these remained for the subset of consented patients). |

#### Protocol Violations

1. One control provider mistakenly received one preference report during the pilot phase. This did not affect the analysis given that pilot participants were a priori excluded from the analyses.
2. Patients with a self-reported history of counseling were initially excluded from being randomized to receive the Depcare tool. To align with our IRB protocol (which did not explicitly exclude those patients) and the system level analyses in which self-reported/outside counseling was not a searchable field, patients who were initially excluded for this reason were re-approached

### Administrative Information

**Scientific Title:** Implementation Science Approach to Enhancing Depression Treatment in integrated primary care settings: The Transform DepCare randomized control trial.

**Public Title:** Implementation Science Approach to Enhancing Depression Treatment in integrated primary care settings: Transform DepCare

**Inclusive Dates of Project:** 09/01/2017- 07/31/2023

**Funding Agency:** Agency for Healthcare Research and Quality (AHRQ) Grant Award Number: R01HS025198

#### Trial Registration Data

**Clinicaltrials.gov**: NCT05085886

**Secondary Identifiers: Columbia University Irving Medical Center: IRB# AAAT6753**

**Primary Sponsor:** Columbia University Irving Medical Center

**Contact for Specific or Public Queries**: ****

**Countries of Recruitment:** USA only

**Health Conditions or problem Studied:** Depression, collaborative care, sustainability

**Interventions:** Randomization of primary care providers to multi-level implementation strategy (intervention arm) versus system-level strategy (enhanced usual care [EUC] arm)

#### Final Eligibility Criteria

Providers

- Inclusion Criteria
  - Primary care providers (PCPs) serving adult patients in one of the five participating clinics

Patients

- Inclusion Criteria
  - Adult patients (18 years or older) with ≥1 completed visit with a PCP in one of the five clinics during the study period (index visit)
  - English or Spanish speaking
  - Elevated depressive symptoms on the day of, or within two weeks prior to, an index visit, defined as:
    - PHQ-9 score ≥ 10
    - OR PHQ-9 score ≥ 5 and GAD-7 score ≥ 10
- Exclusion Criteria
  - Diagnoses of:
    - Coronary heart disease (due to an ongoing study in this population)
    - Dementia
    - Intellectual developmental disorder
    - Severe mental illness (e.g., bipolar disorder, schizophrenia, psychosis)
    - Alcohol/Substance abuse
    - Pregnancy
  - PHQ-9 < 10 or PHQ-9 ≥ 5 with GAD-7 < 10
  - Active enrollment in integrated collaborative care or behavioral health program providing treatment optimization (i.e., completed a visit within the prior 3 months)

#### Status

**Date of Randomization: 8/30/2021**

**Target Sample size 36 providers (actual 44), minimum 252 patients**

**Recruitment Status: Complete**

**Primary outcome:** Percentage of visits for which patients optimize treatment

**Funding:** R01HS025198

### Introduction

**Study Purpose and Rationale**

*Depression is a leading contributor to poor health in the United States.* Depression is common and complicates chronic disease management,^1,2^ reduces quality of life,^3^ and increases functional impairment,^4^ morbidity,^5^ and mortality,^6-8^ particularly in minorities.^9-12^ Depression is ranked the leading cause of years of life lived with disability,^13^ and the third leading cause of loss of quality adjusted life years in older adults.^14^ It is projected that by 2030, depression will be the second leading cause of global burden of disease.^15^ Primary care settings have become the de facto source of depression care in the U.S.^16^ Nonetheless, primary care depression screening, treatment, and remission continue to be suboptimal,^17-19^ resulting in continued patient burden. Though effective treatments for depression exist,^17,18,20^ only 30% of depressed adults who present to their health providers receive guideline-concordant treatment.^21^ Even patients enrolled in accountable care organizations are 24 percent less likely to have their depression or anxiety treated during the following year than patients who remain unenrolled, and see no relative improvements at twelve months in their depression and anxiety symptoms, perhaps due to focus on screening vs. treatment.^22^ Suboptimal depression treatment rates are even higher among low-income and minority patients,^23^ who are more likely to have chronic, disabling depressive symptoms, “no-shows” for treatment sessions, premature treatment termination,^12,24^ and to remain untreated.^10^ The United States Preventive Services Task Force (USPSTF) now recommends depression screening and treatment in primary care settings with systems in place to assure accurate diagnosis, treatment, and follow up.^25^ Only 27% of depressed primary care patients see a significant ≥50% improvement in depressive symptoms.^26^ This rate has remained unchanged for nearly 2 decades^26^ despite increased proportions of primary care visits addressing mental health,^27^ and global improvements in knowledge/comfort among primary care providers (PCP)^28^ as it relates to managing depression though large gaps remain in optimal practices. Some of this may be due to suboptimal focus on treatment optimization, (i.e., referral/treatment intensification, and improved adherence to visits/medications, reach of treat-to-target collaborative care models [CoCM]), particularly in screen-detected historically marginalized populations who are less likely to receive referrals/medications/follow up plans.^29^

*Integrated care models like CoCM are evidence-based approach to managing depressive symptoms in primary care settings.* The primary care medical home (PCMH) model has been heralded as a promising way to improve healthcare in the U.S.; it encompasses comprehensive, patient-centered, coordinated, accessible, high-quality evidence-based care.^30^ Integrated care models like collaborative care for depression, a team-based care management approach, was developed to form a delivery framework for PCMH. The components of CoCM include primary care providers, care management staff trained to provide evidence-based depression care coordination and psychotherapy (by phone or in person), and psychiatric consultants who supervise the primary care team. Other core features include measurement-based care and treat-to-target stepped care, in which treatment is intensified for patients not reaching timely targeted reductions.^16^ Multiple systematic reviews representing over 100 randomized clinical trials (RCTs) establish a strong evidence base for CoCM depression treatment in primary care settings.^16,31,32^ Compared to usual physician care, CoCM significantly improves depressive symptoms by nearly 30*%, halves remission rates*, and increases patient satisfaction, quality of life, productivity, and access to care in both rural and urban settings.^33,34,35-39^ CoCM also appears to impact health disparities, with minorities experiencing greater improvement in depressive symptoms, daily functioning, and receipt of preferred treatment;^40^ unmet appropriate depression care;^41^ and reduced racial discrimination.^42^ Recent studies suggest that collaborative care is effective in older adults, with the minimal number of sessions to be effective being 5 sessions.^43^

*COCM is being widely disseminated, but gaps remain in its sustainability, particularly in minority communities.* CoCM has expanded exponentially across the U.S., facilitated by PCMH initiatives, Accountable Care Organizations, reimbursement for primary care redesign,^44^ and a prior announcements to reimburse for CoCM by the Centers for Medicare and Medicaid Services (CMS).^45^ Multiple systemic reviews have also identified evidence gaps in how best to translate these guidelines into health system practice,^46-48^ particularly in settings serving majority historically marginalized patient populations and those with comorbid conditions.^31^ There have also been recent interest and efforts to improve integration and sustainability of these models with a focus on moving programs along the integration continuum. There have now been a multitude of studies examining the implementation of CC, with varying degrees of success.^49-59^ Thus far, implementation efforts have focused on system-level interventions like financial support/reimbursement codes, quality improvement, training, external technical assistance, and external/internal facilitation.^46,50,51,55,56,60-63^ For example, using a stepped wedge design, the DIAMOND trial found that intensive training/fiscal strategies can improved integrated care implementation processes but not reach or remission rates.^56^ These models are now increasingly reimbursed, and there is consensus that they require intensive implementation support (e.g., training and technical assistance from external experts in collaborative care) to see clinical improvements similar to original trials.^61^

*In real world settings, CoCM sustainability is limited by low patient and PCP engagement.* Even with these resources, patient and PCP engagement remain persistent barriers. In one study of the Office of Mental Health’s Collaborative Care Medicaid Program, a strategy involving technical assistance, fee-for-quality reimbursement, training and quality monitoring, we found that despite steady rates of depression screening, staffing and treatment titration rates and significantly higher contacts/patient and improvement rates clinics enrolled clinics see diminishing reach and fewer patients/ care manager full time equivalent (FTE) over time; key barriers to sustainability noted by respondents included time/resources/personnel, patient engagement and staff/provider engagement.^64^ We estimated that Improving reach of CoCM could reduce mortality by 24% and produce approximately $1300 in annual net savings per patient.^65,66^ PCPs often fail to recognize depression and to refer depressed patients, contributing to suboptimal implementation and low patient engagement rates.^24^ One systematic review found that even when they recognize depression, providers have a poor understanding of CoCm and are often ill equipped to discuss depression.^67^ These communication gaps contribute to depression treatment nonadherence and suboptimal engagement in primary care settings,^68^ including amongst those referred to CoCM.^54,63^ Limited by time, resources, and competing demands, providers are often unable to address barriers to engagement, including misconceptions about depression treatment, stigma, and treatment preference. To date there has been very little focus on treatment optimization behaviors (i.e., intensification, improved reach of treat-to-target CoCM programs).

*Patient-Level strategies may be key to improving engagement.* We hypothesize that population level impact of these programs have been limited by lack of focus on optimization behaviors, including CoCM reach, treatment intensification, combination of medications/therapy. Few have applied a behavior lens to implementation and sustainability efforts. Our systematic review of interventions aimed at improving treatment initiation in depressed primary care patients found that patient preference driven approaches may be impactful.^69^ A cluster randomized controlled trial in 2015 of 117 patients and 301 primary care providers found that the use of a decision aid improved satisfaction with the decision between providers and patients, reducing conflict, and improved patients’ knowledge about treatment options without affecting the duration of the clinical encounter. The decision aid had no effect on clinical outcomes or adherence to drug treatment; the time for the consultation in both intervention and control groups was 40 minutes, so more practicable approaches to shared decision making (SDM) in primary care need to be studied.^70^ While SDM principles are employed by care managers after CoCM enrollment (often long after initial diagnosis), activating patients at the time of diagnosis is key to outcomes.^71^ Computer generated, behavioral theory-informed educational interventions have shown promise in changing patient behaviors.^72,73^ Health information technology/telepsychiatry/teaching workforce about medical conditions have been proposed as methods for improving CoCM implementation in the post-COVID era.^74^ Leveraging SDM facilitators such as multimedia/information technology/automatic triggers and engaging key end users to create a decision tool that automates some of the SDM process (e.g., information exchange) and activates care managers, patients, and providers may be additional approaches to improving CoCM sustainability.^75^

*Theory-driven approaches may be particularly essential to designing effective strategies.* While we hypothesized that a patient-level strategy centered around shared decision-making principles might be essential to improving the sustainability of CoCM, use of theoretical frameworks to select acceptable intervention components is increasingly integral to effective intervention development.^76,77^ One widely used model, Behavior Change Wheel^78^ informed our approach to designing our final strategy for improving patient and PCP engagement behaviors (i.e., optimization behaviors). Given our focus on sustainability, we also identified the Dynamic Sustainability Framework to guide our process, which argues for the need for continuous refinement and improvement of interventions during the sustainability phase, through learning and evaluation, problem solving and ongoing adaptations to interventions to enhance fit between interventions, practice settings/contexts and ecological systems over time.^79^

Our entire implementation strategy development process has been previously described.^80^ Building on previously conducted formative focus groups and stakeholder interviews with CoCM staff, social workers/care managers, psychiatrists, providers, administrators at 8 sites (representing 33 clinics and 1 million socioeconomically and racially/ethnically diverse patients) implementing CoCM across NYS, in **Phase I** of our study we conducted interviews with stakeholders and patients, both those who engaged and didn’t engage in CoCM after referral.^64,81^ We defined the area of sustainability in terms of behavior constructs to identify what needed to change at the system (i.e., care manager/behavioral health provider [BHP]), PCP, and patient levels. We found barriers related to Capability (e.g., knowledge/understanding/language), Opportunity (stigma/culture and environmental context/resources), and Motivation (beliefs about treatment consequences, self-efficacy) at multiple levels. We then identified intervention functions, behavior change techniques and corresponding strategies (based on Expert Recommendations for Implementing Change, ERIC) particularly relevant to sustainability before engaging multidisciplinary stakeholders and end users in rapid cycle, iterative, user-centered design process to operationalize and refine strategy components, including approaches for addressing contextual factors that emerged during COVID19.

The final multi-level implementation strategy included a “*patient-level strategy centered around implementation team/staff-delivered (email/text/in-person based on patient preference) DepCare tool (IR CU19184), which includes enhanced depression and anxiety screening, diagnosis recognition support, patient activation, personalized psychoeducation, patient/care manager videos promoting patient treatment engagement, personalized medication selection support and link to external treatment. The provider-level strategy includes emails of educational/motivational video on CC and optimal management of depression and comorbid anxiety, invitations to problem solving/technical assistance meetings and automatically generated DepCare tool decisional support on individual patient treatment preferences delivered to both the provider and care managers. The clinic level strategy includes quality improvement support and education around valid depression screening as well as local technical support/problem solving for mental health staff/providers co-lead with implementation team members”.* ^80^

In **Phase II**, we will conduct a PCP cluster randomized control trial (RCT) in which we randomize 44 providers to our multi-level strategy (intervention or our system-level strategy (enhanced usual care [EUC]). Patients will be assigned to the intervention or EUC based on their PCP’s allocation. The implementation strategy will be evaluated using the RE-AIM (Reach, Effectiveness, Adoption, Implementation, Maintenance) framework that combines an assessment of maintenance constructs continued use of integrated/CoCM and medications (reach), provider referral/medication intensification behaviors (adoption), and CoCM acceptability/feasibility as well as fidelity to CoCM and strategy (implementation), some of which will be described in a second paper.

**Objectives/Hypotheses**

The overarching goal of this proposal is to conduct an RCT that will rigorously test the effect of our multi-level vs system level strategies, both theory-informed, on patient engagement/optimization behaviors (primary) as well as PCP optimization behaviors (secondary) among patients eligible for CoCM in order to contribute to the sustainability of integrated/CoCM.

**To compare the effectiveness of theory-informed system and multi-level strategies for improving treatment optimization in integrated/CoCM settings serving racial, ethnic, socioeconomically diverse patient population.**

Hypothesis 1:	The proportion of eligible patients who optimize treatment will be higher in the intervention arm vs. EUC arm.

Hypothesis 2: The proportion of patients whose providers optimize treatment will be higher in the intervention arm vs. EUC arm.

Hypothesis 3: The pre- to post- improvements in treatment optimization will be higher in the intervention vs. EUC arm.

**Trial Design**

To accomplish our trial’s objectives, we will randomize PCPs to either the system-level or multi-level strategy both informed by the Behavior Change Wheel/Dynamic Sustainability Framework. All eligible patients of enrolled providers will be included in our analyses. We will recruit and consent a subset of patients, PCPs, and staff/care managers to assess the effectiveness and implementation outcomes of our individual components of our strategies. Outcome assessors that extracted and analyzed data from the EHR will be blinded to allocation. Patients and providers will be unblinded. There will be a 12 month pre-implementation period followed by a 4-6 month implementation period and a 12 month post-implementation period. Patients enrolled in the last month of the post-implementation period will be followed for 6 months.

**Rationale**

**Rationale for Choice of Sites.**

The selection of participating health care organizations was informed by each having: 1) a mature and searchable electronic health record (EHR; 2) an integrated/CoCM program already in place (2) already screening for depression using the Patient Health Questionnaire (PHQ-9); and 4) a clinic director interested in participating. The geographical diversity of these sites will also aid generalizability of results.

**Rationale for Choice of eligibility criteria.**

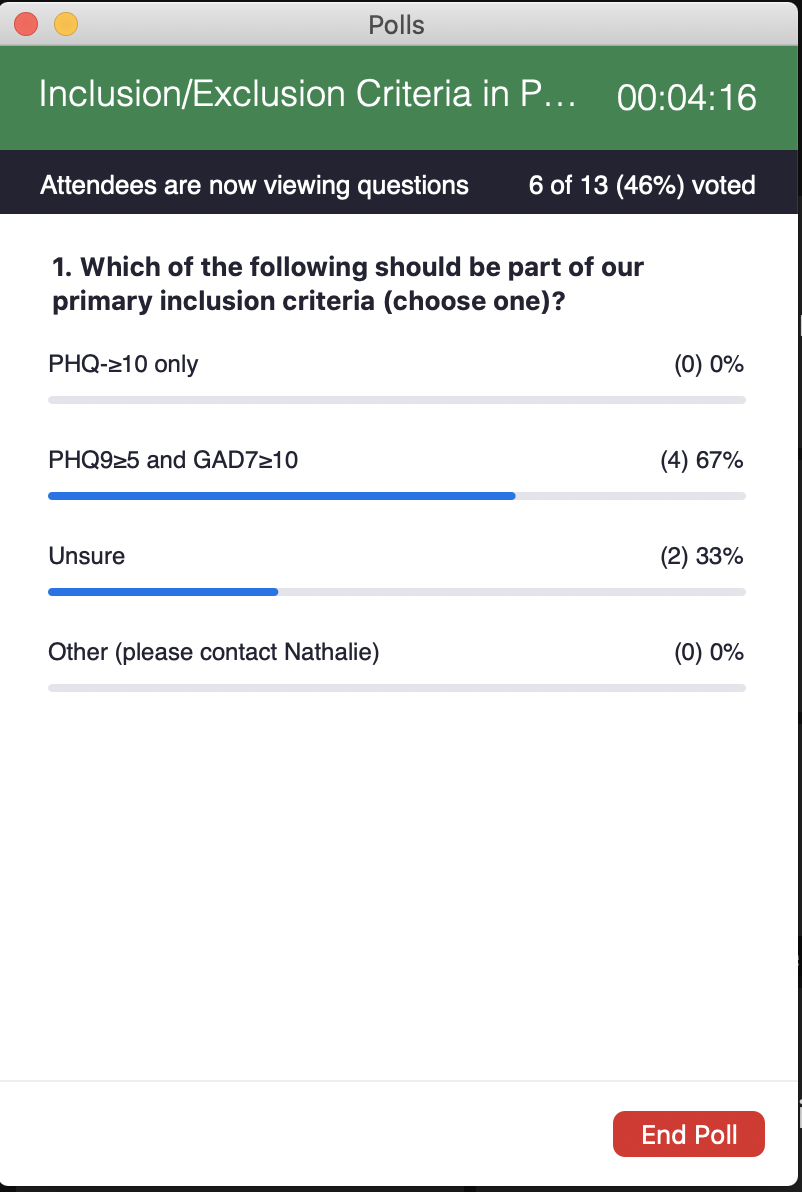
One study found that screening patients without a history of depression yields only 3% positive rates,^82^ suggesting greater utility in including those with a history of depression. Based on our literature review 57% of depressed patients will have chronic depression or fluctuating depressive symptoms throughout their lifetime, which are usually more likely to have more severe depressive and somatic symptoms and greater mental dysfunction at baseline, independent of age, sex, level of education, presence of a chronic disease, and lifetime depression, compared with those who remitted from baseline.^83^ This inclusion criteria was particularly salient to our outcome around optimizing treatment (i.e., increasing doses, ensuring adherence, etc). Few studies have examined ways to optimize depression treatment in these populations, however, and few SDM/patient activation interventions take history of depression and treatment into account. As such we decided to include both those with newly diagnosed depression as well a history of depression.

A key optimization behavior is enrollment in CoCM. As such our wide range of exclusion criteria, including severe mental illness, alcohol abuse, and active participation in behavioral health, CoCM or psychiatry was informed by eligibility criteria for CoCM programs themselves. Our decision to broaden to include anxiety was informed by broadening of eligibility criteria for reimbursement of CoCM across NYS. Along with other eligibility criteria, this decision was also made with input from our advisory board (**Figure 1**). In addition CoCM has demonstrated effectiveness in treating anxiety.^84^ The primary focus on elevated depressive symptoms remained however.

**Rationale for Strategy Components**

Patient-Level: As described in a previous publication,^85^ the tool was iteratively developed based on advisory board feedback, user centered design principles and the Behavior Change Wheel, with the final version demonstrating high usability, and pre-post improvements in decisional conflict, interest in antidepressants, treatment awareness, patient activation and perceived barriers to treatment. Based on advisory feedback, we reduced the focus on SDM to focus on patient activation related to CoCM enrollment and other mental health related behaviors (antidepressant adherence, new antidepressant fill rates, etc). This was also informed by the fact that patient activation has been shown to improve depressive symptoms, depression treatment initiation ^86^ and provider intensification.^87^ Provider-Level: While the BCW revealed that education/training and audit and feedback would be the most feasible behavioral change techniques, research has also shown that patient-targeted along with general practitioner-targeted feedback regarding screening results leads to slightly better improves in terms of depression severity and treatment seeking behaviors.^88,89^ This supported real-time direct feedback regarding depressive symptoms to both patients and providers. System-level: Research has shown that improved depression screening leads to a 1.2% increase rates of systematic screening diagnosis with a screening only trial with an improvement of 5% improvement in treatment (64%-->69%) just by virtue of screening with a 20% increase in odds of treatment pre vs. post implementation.^82^ Given the decay in depression screening in the telemedicine era and following COVID19 disruptions (which markedly reduced screening and reduced the quality of screening), a key clinic level intervention revolved around improved education and support for depression screening. There is also some evidence in the literature to suggest that patients often underreport their depressive symptoms when they are aware they are being screened for depression.^90^ In addition, several studies have suggested that technical assistance, quality improvement, and practice facilitation may be key to improving the uptake of CoCM, further supporting the need for system level strategies.

**Rationale for 4 month follow up period and objective outcomes**

In discussion with statistician and experts we decided to use objective measures to allow for rigorous evaluation of our primary outcome over self-report, particularly given the advantages of Epic. We arrived at an a priori cutoff of 4 months (or nearest visit +/- 1 month for the patient level chart extractions) to account for inherent healthcare system limitations (e.g., long wait times) after consensus with our expert team. We further validated a subset of patients identifying high correlation between self-report and objective Epic data.

### Methods

**Healthcare Setting**

Our study will include 5 primary care clinics with integrated/CoCM programs affiliated with CUIMC: 3 clinics in the Ambulatory Care Network (ACN) of New York Presbyterian serving a predominantly low-income, publicly insured population with substantial numbers of Hispanic and Black or African American patients and 2 Columbia Doctors Primary Care clinics serving a diverse, commercially insured patient population. These clinics are staffed by internal medicine physicians, nurse practitioners and graduate medical education trainees. All clinics had access to core components of integrated care programs:^91^ (1) systematic depression screening with the Patient Health Questionnaire (PHQ-2) and/or General Anxiety Disorder (GAD-2) questionnaire and if positive (score ≥3) a PHQ-9 and/or GAD-7 prior to primary care visits, administrated either verbally by a medical assistant or via the pre-visit e-check-in process and entered into the electronic health record (EHR), Epic (Epic) (2) a behavioral health social worker (BHSW) who provided short-term, measurement-based (PHQ-9) treatment-to-target in collaboration with the PCP (3) use of evidenced based treatment (4) systematic follow up, treatment, and monitoring (5) care coordination (i.e., connection to outside psychiatry, long-term therapy or other services) and (6) systematic consultation and/or program oversight by a psychiatrist/psychologist. All clinics had reimbursement infrastructures for integrated or collaborative care models in place (e.g., Medicare, commercial insurance, and NYS Medicaid reimbursement codes for collaborative care models in the 3 clinics serving the Medicaid population). There was variable access to enhanced external implementation support and varied in program maturity (e.g., ACN clinics received AIMS center expert technical assistance for care managers and launched CoCM 2012; The Columbia Doctors Sites had no external support but had a psychologist lead overseeing the program and launched CoCM in 2021).^61^ All care managers have low caseloads and are available for referrals. In addition to formal CoCM, clinics had varying degrees of access to embedded/integrated behavioral health providers (e.g., psychiatrists, psychologists, nurse practitioners for medication management, and mental health social workers [e.g., to help with coordination of care for more severe cases]. One ACN clinic was reserved for user testing the DepCare Tool.

**Detailed Eligibility Criteria**

As part of the analyses of system vs. multi-level strategy, all PCPs serving adult patients in one of the five participating clinics will be included in our analyses. We will operationalize the following patient level eligibility criteria using EHR query.

Patients can be identified through the EHR if they have ≥1 completed a visit with a PCP in one of the 5 clinics during the relevant study period (referred to as the index visit). Participants are included if they were 18 years or older, English or Spanish speaking, and had elevated depressive symptoms (PHQ9≥10 and/or PHQ9≥5 and GAD≥10) on the day of or during the 2 weeks prior to an index visit (e.g., pre-visit EHR check-ins). Patients are excluded (based on International Classification of Disease -10 codes) if they have a diagnosis of coronary heart disease (due to an ongoing study of depressed coronary heart disease patients), dementia, intellectual development disorder, severe mental illness (i.e., bipolar disorder, schizophrenia, psychosis), substance abuse are pregnant as well as a documented PHQ9 < 10 or PHQ ≥ 5 and GAD <10. Patients are further excluded if they are actively enrolled in a collaborative care or a psychiatry program already providing treatment optimization (i.e., had completed a visit within the prior 3 months). Because psychiatry/CoCM visits can be labeled/misclassified a multitude of ways, any integrated (i.e., embedded in the clinic) behavioral health visit within CUIMC (i.e., psychiatry, psychology, behavioral health, mental health social worker) in the prior 3 months will be excluded. Inclusion/Exclusion criteria were informed by eligibility criteria in collaborative care trials,^92^ and the fact that PCPs might be less likely to make changes if patient is following with a psychiatrist or already enrolled in a collaborative care program with treat-to-target and provider activation protocols. Each visit at which patients had elevated depressive symptoms represents an opportunity to optimize treatment and patients can be eligible at more than 1 visit. All patient visits will be included regardless of whether they are approached and consented for the Depcare Substudy. Our pragmatic approach means that it was possible that patients may be seeing an outside psychiatrist or therapist not captured in our EHR but aligns with our aim to improve optimization regardless of baseline treatment rates. Once an antidepressant script is filled or mental health visit completed, subsequent “index visits” are excluded. Exclusion criteria were based on self-report for the consented DepCare patients.

| **Inclusion Criteria** | **Exclusion Criteria** |
| --- | --- |
| *Inclusion criteria*:   - English- or Spanish-speaking - Age 18 years or older - Elevated PHQ9 >=10 - Elevated PHQ9 >=5 +/- GAD7 ≥10+ | - PHQ9 Screen <10 and/or PHQ9 ≥5 and GAD <10 - History of coronary heart disease (ICD-10 codes: I21, I22, I23, I25). - Pregnancy (O00-O9A) - Schizophrenia/psychosis (F20-F29) or Bipolar Disorder (F30, F31) - History of suicide attempt or self-inflicted injuries - Alcohol/Substance Abuse (F10-F16, F19) - Dementia (Alzheimer’s/Parkinson’s F01-F03, G20-G21, G30-G32, G12-G13 - Cognitive impairment (F70-F89) - Currently under the care or have appointment pending with a psychiatrist or care manager (last visit in last 3 months) (consented patients only) - No access to the internet/smart phone and not planning to attend visit in person (where iPad would be provided to the patient) (consented patients only) - Patients with upcoming psychiatrist/care manager visit in the next 4 months (consented patients only) |

**Intervention and EUC Arms**

The investigators now aim to test this multifaceted implementation strategy, informed by the Behavior Change Wheel, for optimizing treatment amongst patients with elevated depressive symptoms (with or without comorbid anxiety) at CUIMC.

|  | **Experimental Arm** | **EUC Arm** |
| --- | --- | --- |
| **Clinic** | (1) QI Support and education around valid depression and anxiety screening  (2) Local Technical assistance mental health optimization | (1) QI Support and education around valid depression and anxiety screening  (2) Local Technical assistance mental health optimization |
| **Provider** | (3) One-time presentation or video with education and motivational messaging around collaborative care, functionality of the DepCare patient tool and optimal management of depression and comorbid anxiety  (4) 2-4 quality improvement/implementation team meeting on optimizing mental health treatment in primary care and DepCare (i.e., multi-level, multi-component intervention) implementation  (5) Automatically-generated decisional support on individual patient treatment preferences (i.e., for every patient who receives the DepCare patient tool) | (3) Usual Care (social workers are notified of suicidal patients) |
| **Patient** | (6) Patient tool comprised of enhanced depression and anxiety screening (includes option for voice-over questions, point-and-click responses), and for those who screen positive for depressive symptoms (without or without comorbid anxiety), diagnosis recognition support, psycho-education, videos promoting patient engagement in treatment, and personalized medication selection support. | (4) Usual Care (patients are intermittently screened by depression/anxiety screening based on clinic resources or provider indications) |

##### DepCare Intervention Arm (multi-level strategy)

The clinic (administrators, staff, care managers) will receive quality improvement support and education around depression screening as well as local technical assistance for mental health treatment optimization (administrators and champions were to receive technical assistance every 2-3 months in addition to 1:1 input as needed, and activation emails). The cluster of PCPs in the intervention arm will receive education and decisional support for optimizing mental health treatment and access to quality improvement/implementation meetings (including motivational and marketing video embedded in a baseline survey promoting CoCM and treatment optimization; access to a treatment optimization smartphrase in Epic [i.e., Epic tools that allow chunks of text, like “.DepCare” to generate a short user-generated phrase into patient notes], regular emailed motivational newsletters with patient success stories, feedback on screening/optimization rates, reminders/activation around referrals/titrating antidepressants and EHR delivered decision support on individual patient preferences among the subset of eligible patients consenting to receive the Depcare Tool. Eligible patients will receive a tool that facilitates enhanced screening, diagnosis recognition, treatment selection support, psychoeducation, and activation with feedback to PCPs.

##### EUC (system-level strategy)

The clinic (administrators, staff, care managers) will receive quality improvement support and education around depression screening as well as local technical assistance for mental health treatment optimization. The cluster of primary care providers and patients in the active comparator arm will have access to this clinic-level strategy (i.e., the same clinic level intervention as in the DepCare group), but will not receive any provider or patient-level interventions.

**Overview of Study Timepoints and Measures**

The study protocol called for a pre-implementation period that took place from 7/31/2020 to 7/31/2021, an implementation period from 8/1/2021 to 12/31/2021, and a post-implementation period from 1/1/2022 to 12/31/2022. Participants recruited in 12/2022 were followed for 6 months into 6/2023. The key study events were tracked using the Stages of Implementation Completion,^93^ adapted to track implementation strategies in patietns with CoCM already in place. PCPs were randomized on 8/30/2021 in the ACN and 2/7/2022 for ColumbiaDoctors. The pilot study of the DepCare Tool was laucnehd on 8/3/2021 and completed 2/9/2022. The PHQ training started to be disseminated on 1/2021/2022, the first preference report 2/11/2022, The date of the first intervention physician viewed the intervention video (1/13/2022) and the first TA meeting 4/26/2022. The audit/feedback newsletter to providers occurred 11/10/2022. In 12/2022 as study funding ended, ongoing delivery of the Depcare tool was deemed infeasible.

**Primary and Secondary Outcomes**

The *primary outcome* is patient treatment optimization defined as initiating, intensifying, newly adhering (in patients prescribed an antidepressant but with no fills in prior 4 months), or augmenting antidepressants and/or completion of any mental health visit (i.e., integrated/CoCM) during the 4 months following an index visit. Prespecified secondary outcomes include:

(1) the provider taking action to optimize depression treatment at the index visit defined as [a] placing a referral for integrated/CoCM or any mental health provider [b] initiating, intensifying, switching and/or combining antidepressants, providing depression management counseling [i.e., on adherence to treatment regimen]

(2) patient receiving any depression treatment during 4 months post-index visit

(3) among patients initiating an mental health/integrated/CoCM visit within 4 months, those completing ≥2 mental health visits during the 6 months following index visit

(4) among patients initiating an antidepressant within 4 months, those with ≥2 antidepressant fills during 6 months following index visit.

All data will be available via a CUIMC Data Warehouse query for referral orders, patient visits, Patient Health Questionnaire flowsheets, and medication fill dates. To operationalize outcomes, data will be intermittently validated by direct chart review. For example, providers often placed orders for “behavioral health” or “psychiatry” even when they are attempting to refer the patient to CoCM and as such we will include all mental health referrals or visits in our operationalization of “integrated/CoCM” visits and referrals given they are embedded within clinics at CUIMC. We will also validated a subset of antidepressant fill data and confirmed dispense reports. ColumbiaDoctors do not have an extractable orderset for CoCM/Integrated care, which will be conducted via PCP notes and EPIC messages to care managers. Care manager intake sessions will be found in “telephone” encounters. As such referrals and visits will be extracted manually by 2 medically trained abstractors blinded to group assignment. To align with other sites, we will not count referrals to outside therapy/psychiatry or evidence that patients are seeing a counselor outside CUIMC, to align with the fact that this data is not available in data queries in other sites.

For patients randomized to the DepCare substudy, 2 blinded raters will extract patient level metrics, which include baseline treatment and adherence rates, provider behavior at the index visit and subsequent visit, patient engagement rates at 4 and 6 months, depressive symptoms/anxiety at 6 months (**Table 1).** The two raters will enter metrics into REDCAP separately and resolve any conflicts based on consensus. The consensus section will be used for final analyses. Data will be extracted based on ordersets and documented visits, but supplemented using notes (e.g., assessment and plan) to ascertain whether a patient is already in treatment at baseline, nonadherent and/or initiated treatment either within CUIMC (integrated/CoCM) or any mental health treatments (including documentation that patient started treatment outside CUIMC). We also collected the proportion of days covered for anti-depressants at 6 months post index visit.

| **Outcomes on ClinicalTrials.Gov** |  |
| --- | --- |
| Outcome | Description |
| Primary Outcome Measure: Total proportion of patients who initiate or optimize depression treatment | The proportion of patients who initiate or optimize depression treatment, defined as those with at least 1 mental health visit or antidepressant fill during the 4 months following enrollment. Hypothesis 1: this proportion will be greater for patients of providers in the DepCare arm than for the EUC arm [Time Frame: During 4 months post-index visit] |
| Proportion of patients whose providers take action to optimize depression treatment | The proportion of patients whose providers take action to optimize their patients' depression treatment at the index visit, defined as placing a mental health referral for collaborative care or other mental health services; initiating, intensifying, switching and/or combining antidepressant medications; and/or providing depression management counseling [i.e., on adherence to treatment regimen]. Hypothesis 2: this proportion will be greater for the DepCare arm than for the EUC arm [Time Frame: Index Visit] |
| Change in proportion of patients receiving any depression treatment | Change from baseline in the proportion of patients receiving any treatment for depression, defined as filling an antidepressant and/or attending one or more mental health visits during the 4 months following enrollment. Hypothesis 3: This proportion will be greater for the DepCare arm than for the EUC arm. [Time Frame: 4 months pre-Index Visit, 4 months post-index visit] |
| Proportion of patients with at least 2 mental health visits | The proportion of patients with at least 2 mental health visits (i.e., with a mental health specialist). Hypothesis 4: This proportion will be greater for the DepCare arm than for the EUC arm.  [Time Frame: During 6 months post-index visit] |
| Proportion of patients with at least 2 antidepressant fills | The proportion of patients with at least 2 antidepressant fills. Hypothesis 5: This proportion will be greater for the DepCare arm than for the EUC arm.  [Time Frame: During 6 months post-index visit] |
| Mean decisional conflict scale | Mean decisional conflict (measures personal perceptions of uncertainty around choosing among treatment option, 10 items, range 0-100, higher score indicates greater conflict) of patients. Hypothesis 5: the mean decisional conflict will be lower for the DepCare arm than for the EUC arm.  [Time Frame: Baseline] |

**Current Statistical Analysis and Approach**

Patient characteristics at the time of their first index visit are summarized using frequencies and percentages for categorical variables and means [SDs] or medians [IQRs] for quantitative variables. Three closely related hypotheses were tested to evaluate the effectiveness of the intervention (i.e., the multi-level multi-component implementation strategy):

H1:	During the post-implementation period, the proportion of providers’ patients who optimize treatment (attending ≥ 1 integrated/CoCM visit and/or filling a prescription for a new/higher/changed antidepressant) is higher for providers randomized to the intervention arm vs. providers randomized to the enhanced control arm.

H2:	The proportion of patients for whom their provider optimizes treatment is higher for providers randomized to the intervention arm vs. providers randomized to enhanced control arm.

H3:	The improvements in rates of patient and provider (separately) treatment optimization from the pre- implementation period to post-implementation period is higher in the intervention vs. enhanced control arm.

For these analyses predicting patient behavior we estimated a multilevel logistic regression model,^94,95^ where index visits (level 1) are nested within patients (level 2) who are nested within provider (level 3), predicting optimization within 4 months (yes/no), with the Arm (intervention vs EUC) × Period (pre- vs. post-implementation) interaction (4 categories) as the only predictor. Provider (the unit of randomization) and Patient were treated as random effects. Estimates of pre-specified contrasts, corresponding to the above hypotheses, were computed and tested using the Wald F-test (2-tailed, α=0.05). Odds ratios (ORs) and their 95% CIs were obtained by exponentiating the contrast estimates and their 95% CIs. Patients’ outcomes were all analyzed in the arm to which their providers were randomized (**intent-to-treat principle**), regardless of whether they completed the patient-level strategy (DepCare tool). The same approach was used to evaluate the effect of the intervention on secondary patient behavior outcomes, including separate analyses of completing ≥1 CC or psychiatry visit, filling ≥1 new/modified prescription and completing ≥2 behaviors within 6 months of an index visit. For the analyses of provider behavior, we determined the number of patient index visits and proportion of these in which the provider optimized treatment (made a referral to CC or psychiatry and/or wrote a new prescription for or modified the dose of a previously prescribed antidepressant) and used these to estimate a multilevel binomial regression model predicting the proportion of index visits that providers optimized, For these analyses, Provider was treated as a random factor. Because both patient and provider outcomes were based on documentation in the EHR (EPIC), missing data were effectively treated as “non-optimized” behavior, While all visits and medications are well documented in EPIC, we nevertheless note this as a limitation, one that could downwardly bias our estimates of optimization rates but is unlikely to bias estimates of differences between arms or change from pre- to post-implementation periods. We have also performed pre-planned exploratory subgroup analyses to examine whether the effect of the intervention is similar in the Columbia Doctors vs. ACN clinics in order to address potential differences in patient demographics between the 2 settings. We also conducted a sensitivity analysis that excluded PCP champions involved in our system-level intervention (and their patients). Last, we assessed the subgroup of patients who were consented, comparing those who received the Depcare tool vs. usual care. All analyses were performed using the GLIMMIX procedure in SAS 9.4.

Descriptive statistics were used for provider level data. The implementation outcomes were originally administered on a scale of 0-4 and converted to a scale of 1-5 prior to analyses.

**Sample size Calculation**

Our primary aim was to compare the multi-level to the system level implementation strategies. The sample size calculation was originally informed by the observed effect sizes of prior patient-level preference-driven trials on treatment initiation and clinically relevant change in treatment engagement.^96,97^ In our systematic review of interventions shown to improve initiation in depression treatment in primary care settings, patient level interventions (i.e., motivation/activation and treatment preference matching) saw differences in treatment initiation of 15-25% between intervention and usual care arms.^98^ In this review, multi-level and multi-component engagement interventions (vs. enhanced usual care arms) showed effect sizes of 20% (e.g., multicomponent care management and case management interventions).^98^ Based on this evidence, we assumed an effect size of 20% for our primary outcome, patient optimization behavior. Using preliminary pre-implementation data from an ongoing depression screening trial of coronary heart disease patients (i.e., usual care), we found that 20% of patients (10 out of 50) initiated treatment within 3-4 months and that 40% were on treatment at 6 months post index visit. We therefore assumed that those with elevated depressive symptoms in the EUC arm would have a patient optimization rate of **π**=40% and that the patients in the intervention arm would have an optimization rate of **π**=60%, a difference of 20%. In a typical RCT, one without clustering by provider, N=97/arm (with no missing data) would provide 80% power to detect the hypothesized 20% difference between arms (and >80% power to detect a 20% if the patient optimization rate happens to be either higher or lower than **π**=40%). When initially planning this group RCT (GRCT), we anticipated being able to randomize 36 providers. In our preliminary data, we also observed that the intraclass correlation coefficient (ICC), a measure of the degree of clustering by provider, was 0.076. Using the standard power calculation formula for GCRTs,^99^ we determined that 36 providers (18 per arm) with an average of 9 patients with eligible index visits/provider (N=324 patients in total) would provide 80% power to detect the hypothesized 20% difference between arms in patient optimization rates; 17 patients/provider (612 total patients) would provide >90% power]. In the actual study, we were able to randomize 44 providers (605 patients).

The “standard power calculation formula for GCRTs” works well if there is not too much variability from provider to provider in the number of patients/provider. However, 3 providers had no patients with an eligible visit during the post-implementation period and although the number of patients/provider averaged 15.5 among the remaining providers, it ranged from 1 to 110 (median=7, interquartile range: 4 to 18). This certainly reduced the study’s power substantially, compared to a study where each provider had, say, 10-20 patients with an eligible visit. Using the formulas in Hemming et al,^100^ and the observed ICC for providers, 0.031, a sample size of 470 patients with 1 index visit/patient would have been sufficient to provide 80% power to detect the hypothesized effect, and the actual sample size of 605 would provide 88% if we only analyzed patients first eligible index visit. Finally, based on the observed standard errors in the final analysis that included multiple index visits/patient while adjusting for clustering by provider and clustering by patient within provider, we calculate that the study, as conducted, had ~94% power to detect the hypothesized 20% difference between the multi-level multi-component implementation strategy and the enhanced usual care strategy. Thus, the study’s failure to detect the hypothesized effect cannot be attributed to a lack of statistical power.

### Recruitment, Informed Consent and Procedures

To ascertain why our implementation strategy did or did not work, we will recruit and consent a subset of PCPs to administer an exit survey; staff/administrators/BHP to administer pre-post surveys; and a subset of patients to receive the the DepCare tool (based on their PCP’s assignment) followed by a survey. Among the subset of patients consented to participate in the DepCare substudy, two medically-trained abstractors blinded to group assignment independently reviewed the EHR for evidence of patient- and PCP-level optimization. Discrepancies will be resolved through discussion and consensus with a third study team member if needed (none of the cases required a third team member).

**PCPs Recruitment and Procedures.**

We approached clinics through communications with the ACN and Columbia Doctors leadership, specifically the directors of each of the clinics who could decide whether practices should participate in the trial. These health system leaders were provided with details of the implementation strategy prior to agreeing to participate. Altogether 5 clinics agreed to participate. All PCPs in a participating practice will be included in the study. All PCPs will be approached to complete a survey.

PCPs will be contacted via (1) email through the NYP/ACN/Columbia clinic directory with option to learn more about the study by clicking on a link embedded within the email that takes them to an online Qualtrics/REDCap-based information sheet or if preferred undergo verbal consent by phone, via zoom or in-person (2) during staff meetings (i.e. interdisciplinary meeting) or (3) flyers with the QR code of the study. Providers will be contacted via professional networks if they left the clinic. Providers can be approached in person in between clinical sessions as well to complete a paper version of the survey. All providers will be consented.

In Step 1, intervention PCPs will be asked to view a provider education video and complete a brief questionnaire confirming they viewed the video and if they found it acceptable, helpful, and satisfactory followed by a survey. Intervention providers will continue to receive newsletters/updates via email.

In Step 2, both intervention and EUC providers will be asked to provide consent and complete a questionnaire via Qualtrics. Participants will elicit feedback on the acceptability/ satisfaction with and determinants of depression screening and treatment, particularly telepsychiatry and tele-collaborative care, barriers and facilitators, and depression treatment optimization (**Table 2)**.

**Clinic Staff:**

We will recruit administrators, medical assistants, care managers, psychiatrists, psychologists, patient navigators, patient billing coordinators, and patient financial advisors in the clinics. Individuals will be contacted by email through the NYP/ACN/Columbia clinic directory and will have the option to learn more about the study by clicking on a link embedded within the email that takes them to an online RedCap/Qualtrics-based information sheet. Based upon preference, interested staff will also have the option to reach out to a member of the research team to undergo verbal consent by phone, via zoom or in-person. Staff participants will view a depression screening video and complete a study questionnaire via Qualtrics (note: the questions posed in the questionnaire are identical to those administered to provider participants) (**Table 2)**. Staff could be approached in person during clinic hours as well to complete a paper version of the survey. All staff will be consented. In addition, staff who identify themselves as administrators and care managers will also be asked to fill out a separate, specific questionnaire concerning collaborative care and the level of integration at two time points during the study period (at baseline and at 6-months). For both NYP/ACN/Columbia clinic providers and staff, the study will be presented and completed both in-person or on zoom (during non-research related medicine or interdisciplinary problem solving meetings and staff/team “huddles”) as well as via email notification by clinic administrators/directors. The first 30 respondents to the baseline Staff Survey will be entered into a raffle to receive prizes.

**Patient Recruitment Procedures.**

All patients of enrolled PCPs with eligible visits will be included in the analyses. A subset of patients will be approached, screened and consented to elucidate impact of the Depcare tool itself. As part of a quality improvement initiative around improved depression and anxiety screening, clinic administrators and leaders, clinical navigators identified patients with upcoming appointments and approached a random sample of eligible patients (e.g., not receiving treatment from a care manager or psychiatrist) before calling patients to administer the PHQ9/GAD7, which was inputted in the patient’s chart (**Figure 2)**.
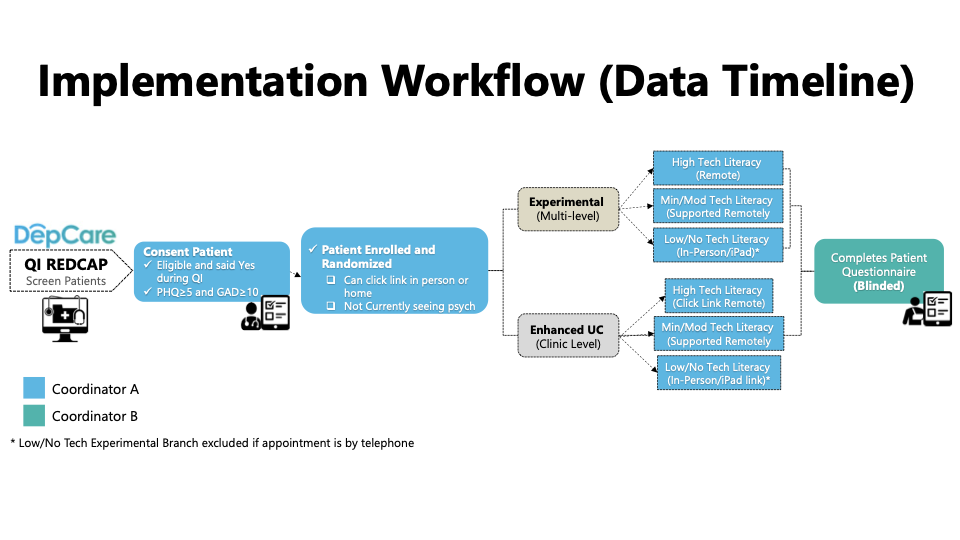

Study activity for patient participants is dependent upon randomization at the provider-level and will include two groups, namely patient participant in Intervention vs. EUC arm. As part of our recruitment approach, the caring clinic provider will approve the enrollment of his/her patient in the study and the first information received by the patient for the purpose of enrollment will be from an individual who has legitimate access to the patient’s medical information. Using the data collected as part of the NYP/ACN/Columbia’s QI initiative, clinic staff will generate a list of patients who a) meet initial study eligibility criteria, b) have the caregiving clinic provider’s approval to approach and c) have been introduced to the study. This list will be sent via an encrypted email message, or via a secure REDCap form to approved research staff on a weekly basis. Using the list provided by clinic staff, members of the research team will access patient medical records under an IRB-approved HIPAA waiver to further screen for exclusion criteria and as applicable, obtain missing data (i.e. a secondary phone number).

For patients who meet study eligibility criteria, research staff will contact them by phone and invite them to participate in our study or coordinate to meet them in-person if the participant prefers. Once the patient is deemed eligible based on a screening survey (i.e., lack of access to the internet and not planning to attend visit in person, not seeing a psychiatrist for a predefined exclusion criteria such as bipolar disorder, suicidality, schizophrenia), the coordinator will receive a notification alerting them to whether the patient is in Intervention or EUC. Patients in both groups will be approached, consented, and enrolled identically (they will have the same consent form and eligibility screening process). Patients who provide consent will undergo a brief screening to confirm eligibility. Those who do not qualify will be informed and thanked for their time.

Patient Participants – EUC: Those patients who qualify will receive a link by email or text (based on preference) thanking them for their time. After they have had their SOC appointment, they will have the option to complete the study questionnaire via RedCap/Qualtrics or via phone or in-person at the patient’s clinic appointment or a scheduled research study visit based on patient preference and coordinator availability.

Patient Participants – Intervention: Those patients who qualify will go on to receive a link to the eSDM tool via text or email, or in-person via an encrypted and registered Columbia University/NYP device. Participants will have the option of completing the DepCare tool (1) on their own (2) with the help of a coordinator by phone or (3) in-person with a device of the participant’s choosing or a study device at their upcoming appointment or at a scheduled research study visit (**Figure 2)**. For all participants in the intervention arm, a summary report will be generated and sent to the patient’s provider and relevant clinic staff regardless of tool completion to ensure providers are notified of their patient’s participation. After they have had their appointment and the opportunity to interact with the tool in the mode that they prefer, participants will have the option to complete the study questionnaire via RedCap or via phone or in-person at the patient’s clinic appointment or a scheduled research study visit based on patient preference and coordinator availability **(Table 1)**. Once consented and randomized, patients will be included in the analyses (e.g., rarely a patient was found to be ineligible e.g., due to serious mental illness during chart extraction of the primary outcome).

| **Table 1. PATIENT – LEVEL SUBSTUDY MEASURES (survey and chart extraction)** | | | |
| --- | --- | --- | --- |
| Measure | Intervention (N=XX) | Control (N=XX) | SOURCE |
| Consent/Medical release | X | X | Survey |
| MRN | X | X | EPIC EHR |
| Patient ID | X | X | Research REDCAP |
| Demographics | X | X | Survey/EHR |
| Eligibility (Not in psychiatry for one of exclusion criteria) | X | X | Survey/EHR |
| On Medication, Taking 90% of time | X | X | Survey |
| EMR Patient contact/update | X | X | HER |
| Patient Health Questionnaire (PHQ) 9^101^ | X | X | QI RedCap |
| Generalized Anxiety Disorder (GAD) 7 | X | X | QI RedCap |
| Type of intervention received (In person/remote/help, brochure)/ Notes | X |  | Research REDCAP |
| Date and completion status of Tool, Brochure, Provider Report (**implementation)** | X |  | Tool Output |
| Preference and Barriers to using DepCare (**implementation)** | X |  | Tool Output |
| Global Provider Trust Question in Primary Care Assessment Survey (PCAS)^102,103^ and Satisfaction with health care for personal or emotional health^33,104^ | X | X | Survey |
| Attitudes around insight into depressive symptoms as well as importance of treating symptoms and intention were adapted from a risk communication and attitudes survey.^105^ (**mechanisms)** | X | X | Survey |
| Decisional conflict Scale (10 item response questionnaire that ascertains the degree to which patients understand their options and any conflicts they have in making decisions about depression treatment with 37.5 indicated decision delay; ≤25 conflict)^102^ (**mechanisms)** | X | X | Survey |
| Depression Treatment Preferences (medications, video or in person counseling, nontraditional options) informed by YouGov survey of MI patients.^106^ | X | X | Survey |
| Current Treatment (meds, therapy) | X | X | Survey |
| Acceptability of both CoCM and DepCare tool based on the four-item validated measure of acceptability of implementation measure- (AIM)^107^ **(implementation)** | X | X | Survey |
| Net promoter Score asks participants “how likely is it that you would recommend this program to a friend on a scale of 0-10”?^108^ **(implementation)** | X | X | Survey |
| Treatment Barriers informed by the National Comorbidity Survey^109^ **(implementation, mechanisms)** | X | X | Survey |
| Physical Activity based on IPAQ | X | X | Survey |
| Loneliness | X | X | Survey |
| EHR extraction of referral, adherence counseling, & medication intensification/initiation | X | X | 6 month Chart Extraction |
| Interim PCP visits Provider Name/Date/Time/Intervention status | X | X | 6 month Chart Extraction |
| EMR extraction of new mental health visit attendance and/or med refill (based on visits and provider/social worker documentation of outside treatment) and Proportion of Days Covered for antidepressants | X | X | 6 month Chart Extraction |

| Table 2. PROVIDERLEVEL SUBSTUDY MEASURES & CLINIC LEVEL PRIMARY ANALYSIS | | | |
| --- | --- | --- | --- |
| Survey | **Intervention (N=XX)** | **EUC (N=XX)** |  |
| Telemedicine Preference, Usability, Satisfaction, Future use | X | X | Survey |
| Depression screening preference | X | X | Survey |
| Determinants/Barriers to telepsychiatry/telecollaborative care informed by the Theoretical Domains Framework and Consolidated Framework for Implementation Research (implementation) | X | X | Survey |
| Acceptability, Feasibility, Appropriateness of Integrated/Collaborative Care^107^ (implementation) | X | X | Survey |
| Net Promoter Score ^108^ (implementation) | X | X | Survey |
| Demographics | X | X | Survey |
| Burnout out: Single-Item Measures of Emotional Exhaustion and Depersonalization in Burnout Assessment^110^ (at least weekly on either emotional exhaustion or depersonalization)^111^ | X | X | Survey |
| PIP is an organizational CoCM self-assessment survey (administrator only)^112^ (implementation/fidelity) | X | X | Survey (administrators psychiatrists/CM only) |
| Organizational readiness for change has been adapted for Implementing Digital Health (median score). Organizational readiness for change is a multi-level, multi-faceted construct. As an organization-level construct, readiness for change refers to organizational members' shared resolve to implement a change (change commitment) and shared belief in their collective capability to do so (change efficacy).^113^  (implementation) | X | X | Survey |
| Types and number of providers who received Training video, in person vs. online (Implementation) | X |  | Qualtrics Survey + SIC Database |
| DepCare Tool feasibility (recruitment, retention, adherence) (implementation) |  |  | Research + QI RedCap |
| Types and number of patients who received DepCare summary report (implementation) | X |  | Tool Database |
| CLINIC LEVEL | | | |
| EHR extraction of visits, flowsheets, referral ordersets, and medication fill data for patient (reach) and provider (adoption) treatment optimization | X | X | EHR |

### Randomization, Allocation and Blinding

**Randomization**

Primary care providers at the 5 participating practices will be stratified by training level (resident, chief/attending, NP). A computer-generated allocation table incorporating permuted block randomization within each stratum (created by the study’s blinded biostatistician [JES]) was used to assign the 36 providers working at these practices in 7/2021 to either the intervention (i.e., multi-level strategy) or enhanced usual care [EUC] (i.e., system-level strategy) arm. Eight new providers (e.g., incoming residents) were randomly allocated to arm at the time they joined a participating practice for a total of 44 providers.

**Blinding**

For the Depcare substudy, we will ensure that the clinical staff/navigator approaches patients without knowing of what intervention arm the PCP is in. Only after enrollment will the coordinator be unblinded to the intervention arm. An outcome assessor blinded to the intervention arm will conduct the outcome assessment. For the primary outcome assessments, clinical individuals outside the team who are blinded to allocation will conduct the chart extractions.

**Ethics and Dissemination**

**Research Ethics Approval**

The study protocol for the consented research components of the study protocol described below was approved by the Institutional Review Board at Columbia University Irving Medical Center (CUIMC). We received approval to analyze data collected as part of quality improvement.

**Informed Consent**

We will obtain informed consent for all staff, patient, and provider surveys including receipt of the Depcare tool. Informed consent will involve a complete description of the study to the participant in clear, easy to understand language. After reviewing and understanding the consent and procedures, those who choose to participate will give their consent to proceed. We obtained a waiver of informed consent for all clinic level analyses to evaluate the impact of our system and multi-level strategies.

**Confidentiality**

As part of the process involved in obtaining informed consent, all participants will be reminded that their responses are confidential and that may refuse to participate in the study or withdraw at any time without explanation and further that such an action will in no way affect their future interactions with their participating Medical Center. To ensure confidentiality, all study-related information will be stored securely at the study sites. When paper records are obtained, those containing names or other personal identifies such as locator forms, medical records, and informed consent forms will be stored separately from study records identified by the participate ID number. Anything linking participant ID to identifying information will be stored in a separate, locked file in the area with limited access. All databases will be secured with pass-word protected access systems. All staff will have all relevant IRB and HIPAA training in the protection of human subject participants (Good Clinical Practice).

**Risks**

This study involves no greater than minimal risk to participants. One possible risk includes the loss of confidentiality of study data. We have a plan in place to protect participant confidentiality.

Additionally, some participants may feel uncomfortable answering study questions about depressive and anxiety symptoms resulting in minor stress. Participants will be reminded that participation is voluntary and that they may refuse to answer any question they choose.

There are no additional anticipated risks associated with the use of the eSDM tool as it has been extensively user tested in other clinic settings and demonstrated high usability and safety when administered to English and Spanish speaking patients across a broad age, gender, race/ethnicity range.

**Benefits**

There is no direct benefit to subjects who participate in this study. The information obtained as a result of this study may be used to inform quality improvement initiatives involving depression screening and thus may benefit future patients.

**Alternatives**

The alternative is not to participate.

**Data and Safety Monitoring**

As this study presents minimal risk to participants, there is no plan for a DSMB.

11.	Sims M, Redmond N, Khodneva Y, Durant RW, Halanych J, Safford MM. Depressive symptoms are associated with incident coronary heart disease or revascularization among blacks but not among whites in the Reasons for Geographical and Racial Differences in Stroke study. *Annals of Epidemiology.*25(6):426-432.

12.	Williams DR, Gonzalez HM, Neighbors H, et al. Prevalence and distribution of major depressive disorder in African Americans, Caribbean blacks, and non-Hispanic whites: results from the National Survey of American Life. *Archives of general psychiatry.* 2007;64(3):305-315.

13.	World Health Organization (WHO). Global Burden of Disease in 2002: data sources, methods and results. World Health Organization (WHO). 2007;Global Programme on Evidence for Health Policy Discussion Paper No. 54.

30.	Agency for Healthcare Research and Quality. Defining the PCMH; <http://pcmh.ahrq.gov/page/defining-pcmh>. . In.

49.	Advancing Integrated Mental Health Solutions (AIMS). In: University of Washington: <https://aims.uw.edu/collaborative-care/implementation-guide>.

50.	Bao Y, Druss BG, Jung H-Y, Chan Y-F, Unützer J. Unpacking Collaborative Care for Depression: Examining Two Essential Tasks for Implementation. *Psychiatric services.* 2015;67(4):418-424.

51.	Chaney EF, Rubenstein LV, Liu CF, et al. Implementing collaborative care for depression treatment in primary care: a cluster randomized evaluation of a quality improvement practice redesign. *Implementation science : IS.* 2011;6:121.

52.	Chaney EF, Rubenstein Lv Fau - Liu C-F, Liu Cf Fau - Yano EM, et al. Implementing collaborative care for depression treatment in primary care: a cluster randomized evaluation of a quality improvement practice redesign

Comparative effectiveness of external vs blended facilitation on collaborative care model implementation in slow-implementer community practices. (1748-5908 (Electronic)).

53.	Eghaneyan BH, Sanchez K, Mitschke DB. Implementation of a collaborative care model for the treatment of depression and anxiety in a community health center: results from a qualitative case study. *J Multidiscip Health.* 2014;7.

54.	Fortney J, Enderle M, McDougall S, et al. Implementation outcomes of evidence-based quality improvement for depression in VA community based outpatient clinics. *Implementation science : IS.* 2012;7:30.

55.	Smith SN, Liebrecht CM, Bauer MS, Kilbourne AM. Comparative effectiveness of external vs blended facilitation on collaborative care model implementation in slow-implementer community practices. *Health services research.* 2020;55(6):954-965.

56.	Solberg LI, Crain AL, Jaeckels N, et al. The DIAMOND initiative: implementing collaborative care for depression in 75 primary care clinics. *Implementation science : IS.* 2013;8:135-135.

57.	Solberg LI, Fischer LR, Wei F, et al. A CQI intervention to change the care of depression: a controlled study. *Effective clinical practice : ECP.* 2001;4(6):239-249.

58.	Tai-Seale M, Kunik ME, Shepherd A, Kirchner J, Gottumukkala A. A case study of early experience with implementation of collaborative care in the veterans health administration. *Popul Health Manag.* 2010;13.

59.	Whitebird RR, Solberg LI, Jaeckels NA, Pietruszewski PB, Hadzic S, Unutzer J. Effective implementation of collaborative care for depression: what is needed? *The American journal of managed care.* 2014;20.

60.	Bao YH, Casalino LP, Ettner SL, Bruce ML, Solberg LI, Unutzer J. Designing payment for collaborative care for depression in primary care. *Health services research.* 2011;46.

69.	Bryant KPJ-K, D; Khodneva, Y; Safford, M; Singer, J; Moise, N. . Age, gender and racial differences in the association between time-varying depressive symptoms and mortality. . SGIM Conference; 2017; Washington, D.C. .

84.	Nafziger, M., & Miller, M. (2013).

Collaborative primary care: Preliminary findings for

depression and anxiety (Doc. No.13-10-3401). Olympia:

Washington State Institute for Public Policy.

85.	Dauber-Decker KL, Serafini MA, Monane R, et al. User-Centered Design of a Preference-Driven Patient Activation Tool for Optimizing Depression Treatment in Integrated Primary Care Settings (The Transform DepCare Study). *Journal of general internal medicine.* 2024.

86.	Moise N, Falzon L, Obi M, et al. Interventions to Increase Depression Treatment Initiation in Primary Care Patients: a Systematic Review. *J Gen Intern Med.* 2018;33(11):1978-1989.

87.	Allen LA, Venechuk G, McIlvennan CK, et al. An Electronically Delivered Patient-Activation Tool for Intensification of Medications for Chronic Heart Failure With Reduced Ejection Fraction: The EPIC-HF Trial. *Circulation.* 2021;143(5):427-437.

88.	Löwe B, Blankenberg S, Wegscheider K, et al. Depression screening with patient-targeted feedback in cardiology: DEPSCREEN-INFO randomised clinical trial. (1472-1465 (Electronic)).

89.	Kohlmann SA-O, Lehmann MA-O, Eisele M, et al. Depression screening using patient-targeted feedback in general practices: study protocol of the German multicentre GET.FEEDBACK.GP randomised controlled trial. (2044-6055 (Electronic)).

90.	Hunt M, Auriemma J Fau - Cashaw ACA, Cashaw AC. Self-report bias and underreporting of depression on the BDI-II. (0022-3891 (Print)).

91.	AIMS Center. Patient-Centered Integrated Behavioral Health Care Principles & Tasks Checklist. In: Washington Uo, ed. *AIMS Center* Seattle, Washington2024.

92.	Unützer J, Katon W, Callahan CM, et al. Collaborative Care Management of Late-Life Depression in the Primary Care SettingA Randomized Controlled Trial. *Jama.* 2002;288(22):2836-2845.

93.	Saldana L, Bennett I, Powers D, et al. Scaling Implementation of Collaborative Care for Depression: Adaptation of the Stages of Implementation Completion (SIC). *Administration and policy in mental health.* 2020;47(2):188-196.

94.	Singer JD, Willett JB. It’s

about time: Using discrete-time survival analysis to study duration and the

timing of events. *Journal of Educational Statistics.* 1993;18:155-195.

95.	Hedeker D, Siddiqui O, Hu FB. Random-effects regression analysis of correlated grouped-time survival data. *Stat Methods Med Res.* 2000;9(2):161-179.

96.	Kravitz RL, Franks P, Feldman MD, et al. Patient engagement programs for recognition and initial treatment of depression in primary care: a randomized trial. *Jama.* 2013;310(17):1818-1828.

97.	Davidson KW, Bigger JT, Burg MM, et al. Centralized, Stepped, Patient Preference-Based Treatment for Patients With Post-Acute Coronary Syndrome Depression: CODIACS Vanguard Randomized Controlled Trial. *JAMA internal medicine.* 2013:1-8.

98.	Moise N, Falzon L, Obi M, et al. Interventions to Increase Depression Treatment Initiation in Primary Care Patients: a Systematic Review. *Journal of general internal medicine.* 2018;33(11):1978-1989.

99.	Murray DM: The Design and Analysis of Group-Randomized Trials. London: Oxford, University Press; 1998.
