## Supplemental Material 2 for "Comparing implementation strategies for optimizing depression care: A randomized control trial"

**Supplementary Material 2**

**Title:** Comparative effectiveness of multi-level vs. system-level interventions for optimizing depression treatment in integrated primary care settings: The Transform DepCare randomized control trial.

**Authors** Nathalie Moise, Maria Serafini, Danielle Rome, Jennifer Mizquiri Barbecho, Kirali Genao, Siqin Ye, Andrea Duran, Joseph E. Schwartz

**Running title: Sustaining integrated care programs**

eTable 1. Number of visits per provider for EUC and Intervention arms, Pre and Post implementation.

| Period | Condition | 1 visit | 2 visits | 3 visits | >=4 visits | Total |
| --- | --- | --- | --- | --- | --- | --- |
| **Pre-implementation** | EUC | 146 (75%) | 34 (17%) | 8 (4%) | 7(4%) | 195 |
|  | DepCare | 158 (78%) | 38 (19%) | 5 (2%) | 2(1%) | 203 |
|  | Total | 304 (76%) | 72(18%) | 13(3%) | 9(2%) | 398 |
| **Post-implementation** | EUC | 240(81%) | 41(14%) | 11(4%) | 3(1%) | 295 |
|  | DepCare | 277(85%) | 39(12%) | 6(2%) | 3(1%) | 325 |
|  | Total | 517(83%) | 80(13%) | 17(3%) | 6(1%) | 620 |

| **eTable 2: Characteristics of sample, by group assignment and period** | | | | | | | | | | |
| --- | --- | --- | --- | --- | --- | --- | --- | --- | --- | --- |
|  | Pre-intervention | | Post-intervention | | p-values | | | | | |
| Characteristics (n, %) | A. Enhanced Usual Control (N=195) | B. DepCare Intervention (N=203) | C. Enhanced Usual Control (N=295) | D. DepCare Intervention (N=325) | B vs A | D vs C | C vs A | D vs B | 2-way interaction | A vs B vs C vs D |
| Age, mean (SD) | 47.9 (18.5) | 49.9 (16.4) | 46.2 (16.7) | 49.0 (17.6) | 0.23 | 0.046 | 0.30 | 0.54 | 0.75 | 0.084 |
| Female | 162, 83% | 160, 79% | 238, 81% | 262, 81% | 0.28 | 0.95 | 0.50 | 0.57 | 0.38 | 0.76 |
| Male | 33, 17% | 43, 21% | 57, 19% | 62, 19% |  |  |  |  |  |  |
| Nonbinary* | 0 (0%) | 0 (0%) | 0 (0%) | 1, <1% |  |  |  |  |  |  |
| White | 40, 21% | 48, 24% | 98, 33% | 85, 26% | 0.35 | 0.023 | 0.008 | 0.78 | 0.19 | 0.012 |
| Black | 28, 14% | 41, 20% | 36, 12% | 56, 17% |  |  |  |  |  |  |
| Asian* | 5, 3% | 2, 1% | 13, 4% | 13, 4% |  |  |  |  |  |  |
| American Indian/Alaskan* | 1, <1% | 1, <1% | 2, <1% | 0 (0%) |  |  |  |  |  |  |
| Native Hawaiian or Pacific Islander* | 0 (0%) | 0 (0%) | 0 (0%) | 0 (0%) |  |  |  |  |  |  |
| Combination/multi-racial | 86, 44% | 80, 39% | 93, 32% | 127, 39% |  |  |  |  |  |  |
| Declined to respond | 35, 18% | 31, 15% | 53, 18% | 44, 14% |  |  |  |  |  |  |
| Non-Hispanic | 47, 24% | 50, 25% | 95, 32% | 114, 35% | 0.96 | 0.70 | 0.03 | 0.008 | 0.79 | 0.008 |
| Hispanic | 118, 61% | 127, 63% | 149, 51% | 167, 51% |  |  |  |  |  |  |
| Declined/Unknown* | 30, 15% | 26, 13% | 51, 17% | 44, 14% |  |  |  |  |  |  |
| Preferred Language English | 116, 59% | 115, 57% | 200, 68% | 191, 59% | 0.59 | 0.024 | 0.035 | 0.47 | 0.30 | 0.030 |
| Preferred Language Spanish | 74, 38% | 82, 40% | 84, 28% | 119, 37% |  |  |  |  |  |  |
| Other* | 4, 2% | 5, 2% | 10, 3% | 14, 4% |  |  |  |  |  |  |
| Declined/Unknown* | 1, <1% | 1, <1% | 1, <1% | 1, <1% |  |  |  |  |  |  |
| Antidepressant at baseline | 63, 32% | 82, 40% | 127, 43% | 131, 40% | 0.09 | 0.49 | 0.02 | 0.98 | 0.08 | 0.12 |
| Ambulatory Care Network (Medicaid) | 137, 70% | 145, 71% | 133, 45% | 175, 54% | 0.80 | 0.03 | 0.001 | 0.001 | 0.28 | 0.001 |
| Columbia Doctors (Commercial Insurance) | 58, 30% | 58, 29% | 162, 55% | 150, 46% |  |  |  |  |  |  |
| * Category ignored in computation of p-values comparing four groups | | | | | | | | | | |

| **e-Table 3. Pre-Post Optimization Behaviors in Enhanced Usual Care vs. DepCare Intervention Arms** | | | | | | | | | | | | |
| --- | --- | --- | --- | --- | --- | --- | --- | --- | --- | --- | --- | --- |
|  | **Pre-Post Enhanced Usual Care**  **Yes: No (%)** | | | | |  | | **Pre-Post DepCare Intervention** | | | | |
| **Patient Optimization** | **Post** | **Pre** | **OR (95% CI)** | **p-value** | | | **Post** | | **Pre** | **OR (95% CI)** | **p-value** | **Differential p value** |
| ≥1 Fill and/or integrated/CoCM visit at 4 months | 136:167 (44.9%) | 69:158 (30.4%) | 1.93 (1.31, 2.85) | **0.001** | | | 132:206 (39.1%) | | 73:170 (30.0%) | 1.37 (0.94, 2.00) | 0.10 | 0.22 |
| ≥2 Fills and/or visits at 6 months | 113:190 (37.3%) | 58:169 (25.6%) | 1.77 (1.19, 2.64) | **0.005** | | | 106 :232 (31.4%) | | 63:180 (25.9%) | 1.23 (0.83, 1.81) | 0.29 | 0.20 |
| ≥1 Fill at 4 months | 117:215 (35.2%) | 58:171 (25.3%) | 1.63 (1.10, 2.40) | **0.015** | | | 99:266 (27.1%) | | 59:188 (23.9%) | 1.05 (0.71, 1.55) | 0.81 | 0.12 |
| ≥2 Fills at 6 months | 93:239 (28.0%) | 45:184 (19.7%) | 1.61 (1.06, 2.44) | **0.026** | | | 82:283 (22.5%) | | 51:196 (20.6%) | 1.05 (0.70, 1.58) | 0.83 | 0.15 |
| ≥1 Integrated/CoCM visit at 4 months | 67:299 (18.3%) | 28:239 (10.5%) | 2.02 (1.24, 3.30) | **0.005** | | | 62:322 (16.1%) | | 24:229 (9.5%) | 1.86 (1.11, 3.13) | **0.02** | 0.82 |
| ≥2 Integrated Care visits at 6 months | 56:310 (15.3%) | 24:243 (9.0%) | 1.91 (1.14, 3.22) | **0.014** | | | 39:345 (10.2%) | | 19:234 (7.5%) | 1.39 (0.77, 2.51) | 0.28 | 0.42 |
| **Provider Optimization** |  |  |  | |  | |  | |  |  |  |  |
| Med Intensification and/or referral | 140:164 (46.1%) | 77:141 (35.3%) | 1.49 (1.00, 2.22) | | **0.05** | | 135:198 (40.5%) | | 70:161 (30.3%) | 1.24 (0.83, 1.86) | 0.28 | 0.52 |
| Med Intensification | 87:250 (25.8%) | 52:175 (22.9%) | 1.09 (0.71, 1.69) | | **0.69** | | 69:294 (19.0%) | | 37:202 (15.5%) | 1.02 (0.63, 1.66) | 0.92 | 0.84 |
| Integrated/CocM Referral | 98:245 (28.6%) | 42:216 (16.3%) | 1.84 (1.25, 2.70) | | **0.003** | | 98:274 (26.3%) | | 41:205 (16.7%) | 1.36 (0.92, 2.01) | 0.12 | 0.28 |

PCP Primary care Provider

| **e-Table 4. Patient Optimization Behaviors in Enhanced Usual Care vs. DepCare Intervention Arms in Columbia Doctors and Ambulatory Care Network Clinics** | | | | | | | | | |
| --- | --- | --- | --- | --- | --- | --- | --- | --- | --- |
|  | **Ambulatory Care Network** | | | | | **Columbia Doctors** | | | |
| **Patient Optimization** | **Enhanced Usual Care** Yes: No (%) | **Intervention**  Yes: No (%) | **OR (95% CI)** | **p-value**^a^ | | **Enhanced Usual Care**  Yes: No (%) | **Intervention**  Yes: No (%) | **OR (95% CI)** | **p-value** |
| Post | 56:77 (42.1%) | 66:116 (36.3%) | 0.81 (0.47,1.39) | | 0.43 | 80:90 (47.1%) | 66:90 (42.3%) | 0.91 (0.29, 2.84) | 0.87 |
| Pre | 49:105 (31.8%) | 49:131 (27.2%) | Post vs. Pre  1.57 (0.94, 2.61) | | 0.09 | 20:53 (27.4%) | 24:39 (38.1%) | Post vs. Pre  2.75 (1.44, 5.26) | 0.003 |

^a^P value for Differential change by assignment: ACN p= 0.81;Columbia Doctors p=0.09

| **e-Table 5. Patient Optimization Behaviors in Enhanced Usual Care vs. DepCare Intervention Arms excluding primary care champions (Yes:No [%]** | | | | | |
| --- | --- | --- | --- | --- | --- |
| **Patient Optimization** | **Enhanced Usual Care**  Yes: No (%) | **Intervention**  Yes: No (%) | **OR (95% CI)** | **p-value** | |
| ≥1 Fill and/or integrated/ CoCM visit at 4 months | 69:100 (40.8%) | 124:191 (39.4%) | 0.79 (0.47,1.33) | | 0.47 |

**eFigure 1. Consort Flow Diagram Pre-Intervention: Enrollment, Eligibility, and analysis of pre-intervention participants of the Transform DepCare Study**

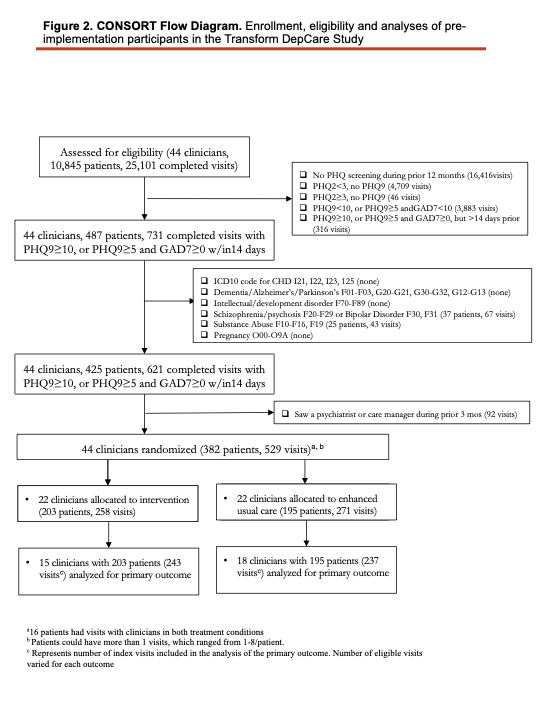

**eFigure 2. Flow diagram for the DepCare Sub-study**

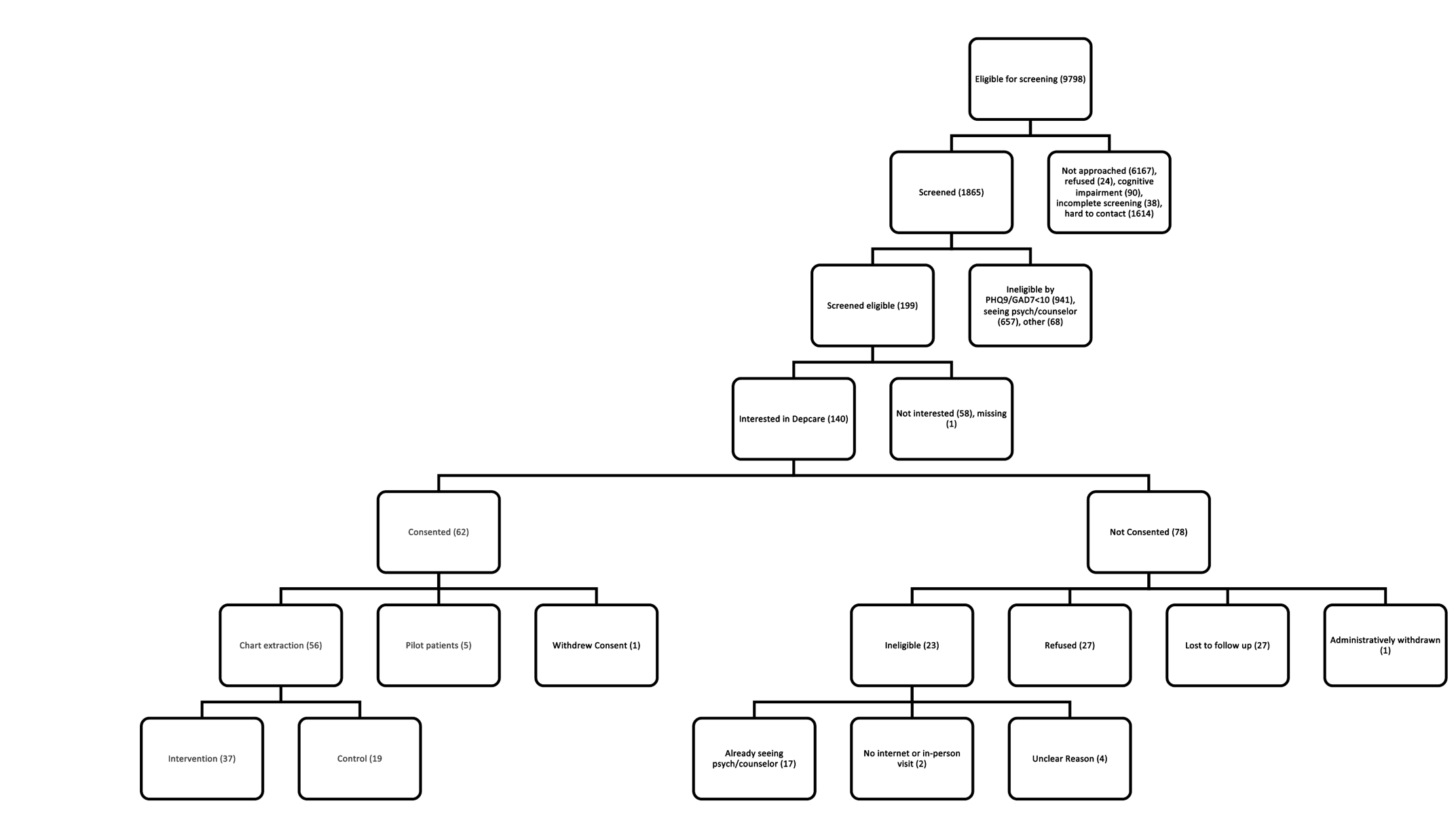

**eFigure 3. Primary Care Physician Collaborative Care Implementation Outcomes in the Enhanced Usual Care Arm compared to the Intervention Arm (Range 1-5).**

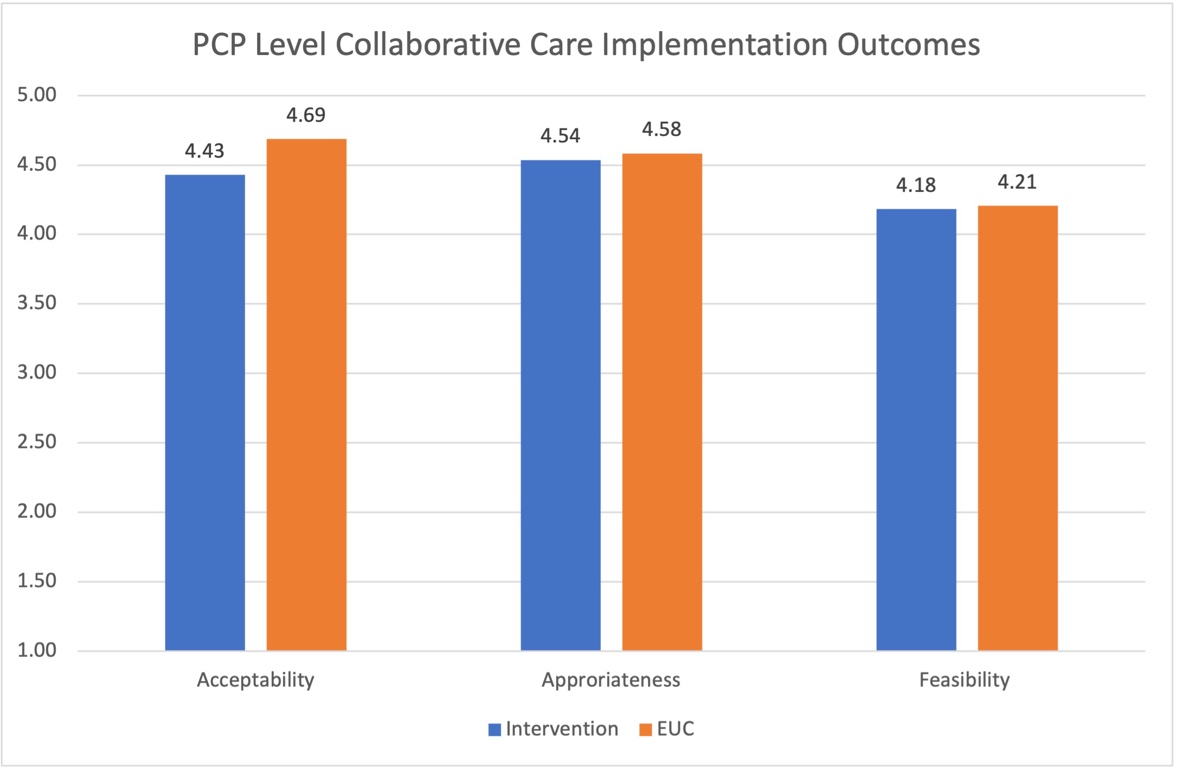
